# Intracranial pressure waveform estimation using extracranial signals with deep learning

**DOI:** 10.64898/2026.06.07.26354958

**Authors:** Ansh Goyal, Veet Zaveri, Carl Harris, Robert D Stevens

## Abstract

Continuous intracranial pressure (ICP) monitoring requires invasive instrumentation and reaches only a subset of the critically ill patients who may need it. We tested whether deep learning models trained on routinely acquired extracranial signals can estimate ICP waveforms with clinically motivated point-estimate accuracy. Five architectures were trained on arterial blood pressure (ABP), photoplethysmography (PPG), and electrocardiography (ECG) waveforms, using invasive intraparenchymal ICP as ground truth, and compared across two fusion strategies and three training objectives. Models were externally validated on the independent MIMIC-III (Medical Information Mart for Intensive Care III) Waveform Database. Performance was assessed by mean absolute error (MAE) and Pearson correlation (r). Across 158 critically ill adults (∼5,322 hours) from two institutions, external MAE of configurations selected using internal validation data only ranged from 4.266 to 5.899 mmHg and r from 0.585 to 0.719; three of five architectures (multiscale encoder-decoder, long-memory recurrent, temporal convolutional) fell below a clinically motivated 5 mmHg benchmark. Unlike prior work validated within single-site cohorts, these models generalized across institutions at the level of mean pressure and pulsatile trend, but did not reproduce beat-level peak morphology. This work extends prior research through large-cohort, cross-institutional validation and motivates evaluation across broader populations and ICP ranges.

## Introduction

Intracranial hypertension is a major driver of secondary neurological injury and adverse outcome in patients with acute brain injury. As such, monitoring intracranial pressure (ICP) in critically ill patients is an important measure to ensure adverse hypertensive events are handled in a timely manner. However, the clinical standard for ICP assessment is intraparenchymal or intraventricular monitoring, which carries procedural risk and is generally limited to settings with advanced surgical and critical care infrastructure [1]. Ventricular catheter-based ICP monitoring carries a clinically meaningful risk of cerebrospinal fluid (CSF) infection and ventriculitis. Reported rates vary widely with case-mix and infection definition (pooled ∼8–13%, range 1.9–36%), and longer catheter dwell time is among the most consistently identified risk factors [2]. Many patients who are at risk for elevated ICP do not receive appropriate monitoring either because they find themselves in a lower-resource setting or because invasive monitoring is reserved for a narrow subset of patients even in well-resourced centers [3].

Research on non-invasive ICP assessment includes indirect ICP surrogates and physiological signal-derived estimation methods [4]. Surrogate techniques such as optic nerve sheath diameter ultrasonography, pupillometry, retinal imaging, and near-infrared spectroscopy-based optical sensing [5] can help identify discrete instances of intracranial hypertension at a specific point in time; they do not provide continuous pulsatile ICP waveforms, are operator-dependent, and can be difficult to apply consistently in critically ill patients [1, 6, 7]. Physiological modeling approaches include frequency-domain methods such as cerebral blood flow velocity (CBFV) acquired via transcranial Doppler (TCD) ultrasonography, a technique which depends on operator expertise and a favorable transcranial insonation window [8, 9]. A recent narrative review identified continuous waveform reconstruction as an unmet need in non-invasive ICP monitoring, noting that existing methods primarily detect whether a predefined threshold is exceeded rather than providing truly continuous measurements, and that current machine learning approaches remain limited by small patient cohorts and dependence on signals not universally available at the bedside [10].

More recently, deep learning has been applied to recover ICP-related information directly from routinely collected physiologic waveforms. Detection and forecasting studies have shown that intracranial hypertension events can be predicted from extracranial signals such as arterial blood pressure (ABP), electrocardiography (ECG), and photoplethysmography (PPG), supporting the idea that these waveforms contain ICP-relevant information [11, 12, 13]. Waveform reconstruction studies have demonstrated that segment-level ICP waveforms can be estimated from ABP alone using encoder-decoder architectures [14] and that domain-adversarial training can improve non-invasive ICP waveform derivation from multimodal hemodynamic inputs [15]. Our group recently reported that algorithmic prediction of the ICP waveform from a discrete set of extracranial continuous physiologic signals yields accuracies that are higher than most other extracranial techniques and on par with invasive techniques like intraparenchymal monitoring [16]. Additionally, a recent study trained and validated an ABP and ECG based model on the Rutgers based Cerebral Haemodynamic Autoregulatory Information System (CHARIS) dataset to remarkable results [17]. However, all of these studies are limited by small cohorts, absence of external validation, and, in some cases, dependence on inputs such as TCD-derived CBFV.

Our objective in this work is to develop a novel, continuous, extracranial signal-based ICP waveform reconstruction approach by training different deep learning models with continuous waveform data from signals which are routinely acquired in virtually any intensive care unit (ICU) (ECG, ABP, and PPG). Models are trained on a large institutional dataset (Precision Medicine Analytics Platform [PMAP], Johns Hopkins) and externally validated on the Medical Information Mart for Intensive Care III (MIMIC-III) Waveform Database using invasively acquired ICP waveform as the ground truth label. We compare five sequence-to-sequence architectures, two multimodal fusion strategies, and three training objectives, one of which up-weights elevated-ICP segments to address the severe underrepresentation of hypertensive ICP segments in clinical data. We hypothesized that continuous ICP waveform reconstruction can be achieved at an accuracy within a clinically motivated mean absolute error (MAE) benchmark, and that the model’s learned representations generalize across hospitals to previously unseen cohorts.

## Results

**Fig. 1** provides a study design overview. We analyzed 5,322 hours of continuous ICU waveform data from a total of 158 patients across two institutions (47 training/internal validation patients, 111 external validation patients). Clinical characteristics of the study population are summarized in **Table 1**. The PMAP and MIMIC-III cohorts were not explicitly matched for illness severity and exhibited differences in their clinical profiles. The PMAP cohort had a higher median Elixhauser Van Walraven score than the MIMIC-III cohort (15 [interquartile range (IQR) 11–22] versus 6 [IQR 3–12]), indicating a greater overall comorbidity burden. In-hospital mortality was also numerically higher in PMAP (29.8% versus 22.5%). The cohorts also differed in diagnostic composition, with a greater proportion of brain tumor or mass diagnoses in PMAP and a greater proportion of other intracranial hemorrhage diagnoses in MIMIC-III. Taken together, these findings indicate that the cohorts were not directly severity matched and may have differed in overall clinical complexity. However, because the Elixhauser score and hospital length of stay do not directly measure the severity of acute neurologic injury, and diagnosis-specific severity measures were not consistently available across both datasets, we could not determine whether patients with the same diagnosis had comparable neurologic disease severity.

**Fig. 1.**
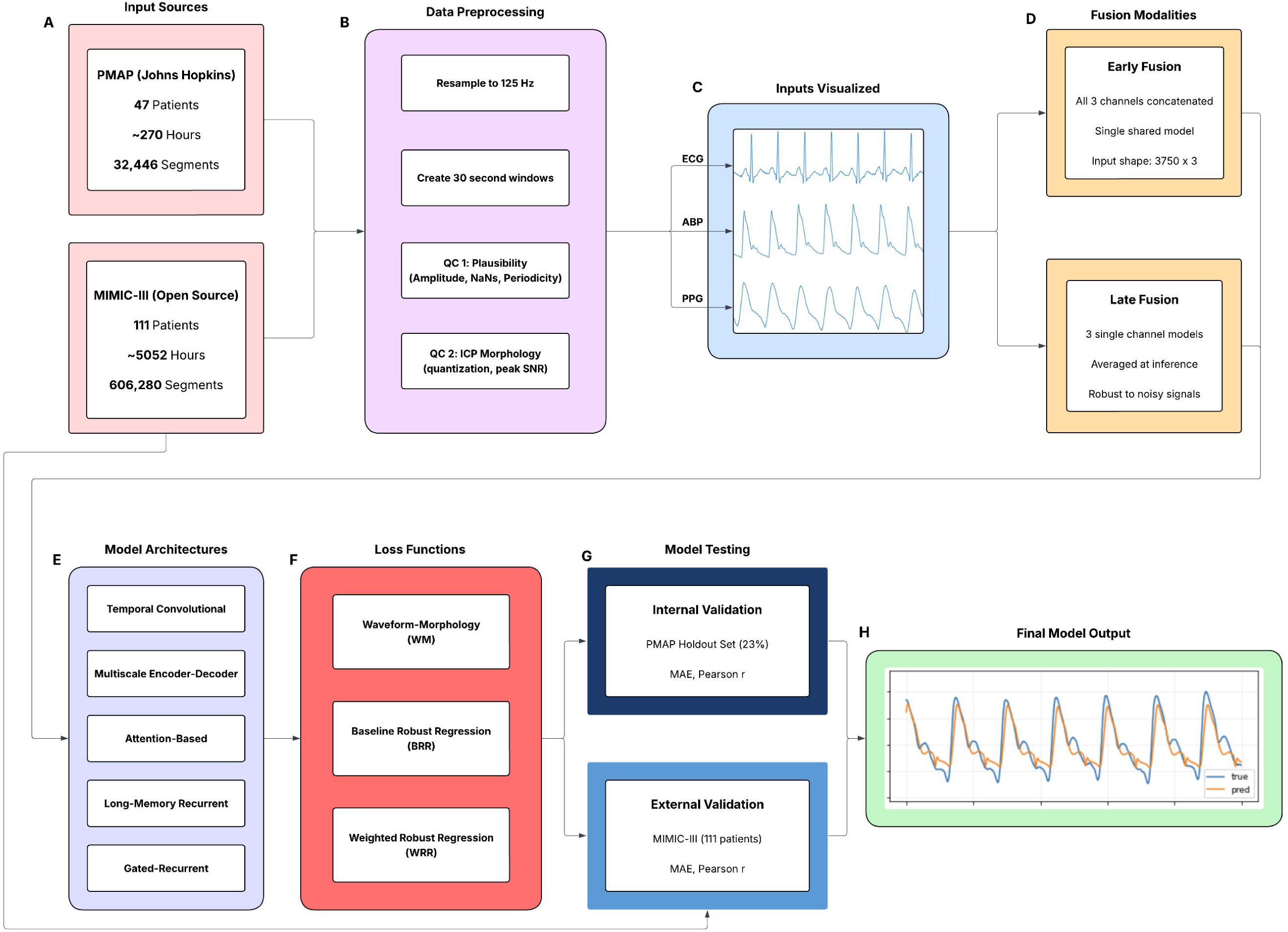
Study design and analysis pipeline. a) Data was collected from PMAP and MIMIC-III for internal and external validation, respectively. b) All data was sent through a standard processing workflow. c) The model inputs contained all 3 extracranial signals. d) Two fusion methods were tested, one that concatenated all inputs in training, the other that averaged individual predictions at inference. e) 5 model architectures were tested. f) Each architecture was tested with 3 loss functions. g) Every model configuration was validated internally (PMAP) and externally (MIMIC). h) The final model predictions were plotted alongside the true waveforms for visual inspection.

**Table 1.** Demographic and clinical characteristics of the PMAP training cohort and MIMIC-III external validation cohort.

| **Characteristic** | **PMAP, n = 47** | **MIMIC-III, n = 111** |
|---|---|---|
| **Age group, n (%)** |  |  |
| 18–39 years | 10 (21.3%) | 12 (10.8%) |
| 40–59 years | 19 (40.4%) | 41 (36.9%) |
| 60–79 years | 14 (29.8%) | 47 (42.3%) |
| ≥80 years | 4 (8.5%) | 11 (9.9%) |
| **Sex, n (%)** |  |  |
| Male | 22 (46.8%) | 46 (41.4%) |
| Female | 25 (53.2%) | 65 (58.6%) |
| **Elixhauser Score, median [IQR]** | 15 [11, 22] | 6 [3, 12] |
| **Primary diagnosis, n (%)** |  |  |
| Subarachnoid hemorrhage/aneurysm | 13 (27.7%) | 35 (31.5%) |
| Brain tumor/mass/neoplasm | 11 (23.4%) | 9 (8.1%) |
| Other intracranial hemorrhage/bleed | 8 (17.0%) | 38 (34.2%) |
| Trauma-related diagnosis | 3 (6.4%) | 5 (4.5%) |
| Other neurologic/neurosurgical diagnosis | 4 (8.5%) | 9 (8.1%) |
| Other medical/non-neurologic diagnosis | 8 (17.0%) | 15 (13.5%) |
| **Hospital length of stay range, days** | 2.6 - 223.4 | 1.3 - 61.9 |
| **In-hospital mortality, n (%)** | 14 (29.8%) | 25 (22.5%) |

All performance metrics reported in the main text correspond to the MIMIC-III Matched Subset cohort (111 patients, 606,280 segments), for which linked clinical demographics are available.

### Internal Validation vs. External Validation Performance

Internal validation was performed using a segment-level split of the PMAP cohort, whereas external validation was performed using the entirely held-out MIMIC-III cohort. **Table 2** shows the external performance of the configuration selected for each architecture using the balanced score computed on PMAP internal-validation metrics alone, so that no external (MIMIC-III) information influenced model selection (see Methods). External MAE ranged from 4.266 to 5.899 mmHg. Point estimates were within the clinically motivated <5 mmHg benchmark for three of five architectures (multiscale encoder-decoder 4.266, long-memory recurrent 4.637, and temporal convolutional 4.847 mmHg), whereas the attention-based (5.285 mmHg) and gated recurrent (5.899 mmHg) models exceeded it. We therefore treat the multiscale encoder-decoder, long-memory recurrent, and temporal convolutional models as the principal architectures, with the attention-based and gated recurrent models reported for completeness. Additionally, external Pearson correlations ranged from 0.585 to 0.719. The attention-based model achieved the highest external correlation (r = 0.719), followed by the temporal convolutional model (r = 0.663), whereas the multiscale encoder-decoder achieved the lowest external MAE. For comparison, **Table 3** reports the configurations that would have been chosen had selection used the external MIMIC-III metrics. Because this selection used the evaluation data, these results represent an optimistic upper bound (external MAE 4.276–4.946 mmHg, r 0.599–0.722, all five models clear the <5 mmHg benchmark). The cost of selecting on internal metrics alone was largest for the gated recurrent model (external MAE higher by 1.623 mmHg, 95% CI 1.121 to 2.148) and the temporal convolutional model (0.489 mmHg, 95% CI 0.146 to 0.852). In contrast, internal selection of the multiscale encoder-decoder yielded a lower external MAE (0.228 mmHg lower, 95% CI 0.033 to 0.452) but a lower correlation (r lower by 0.135, 95% CI 0.094 to 0.168). Representative reconstructed and reference ICP waveforms for each architecture are shown for a pathologically altered segment (Fig. 3) and a normal, P1-dominant segment (Fig. 4).

**Table 2.**
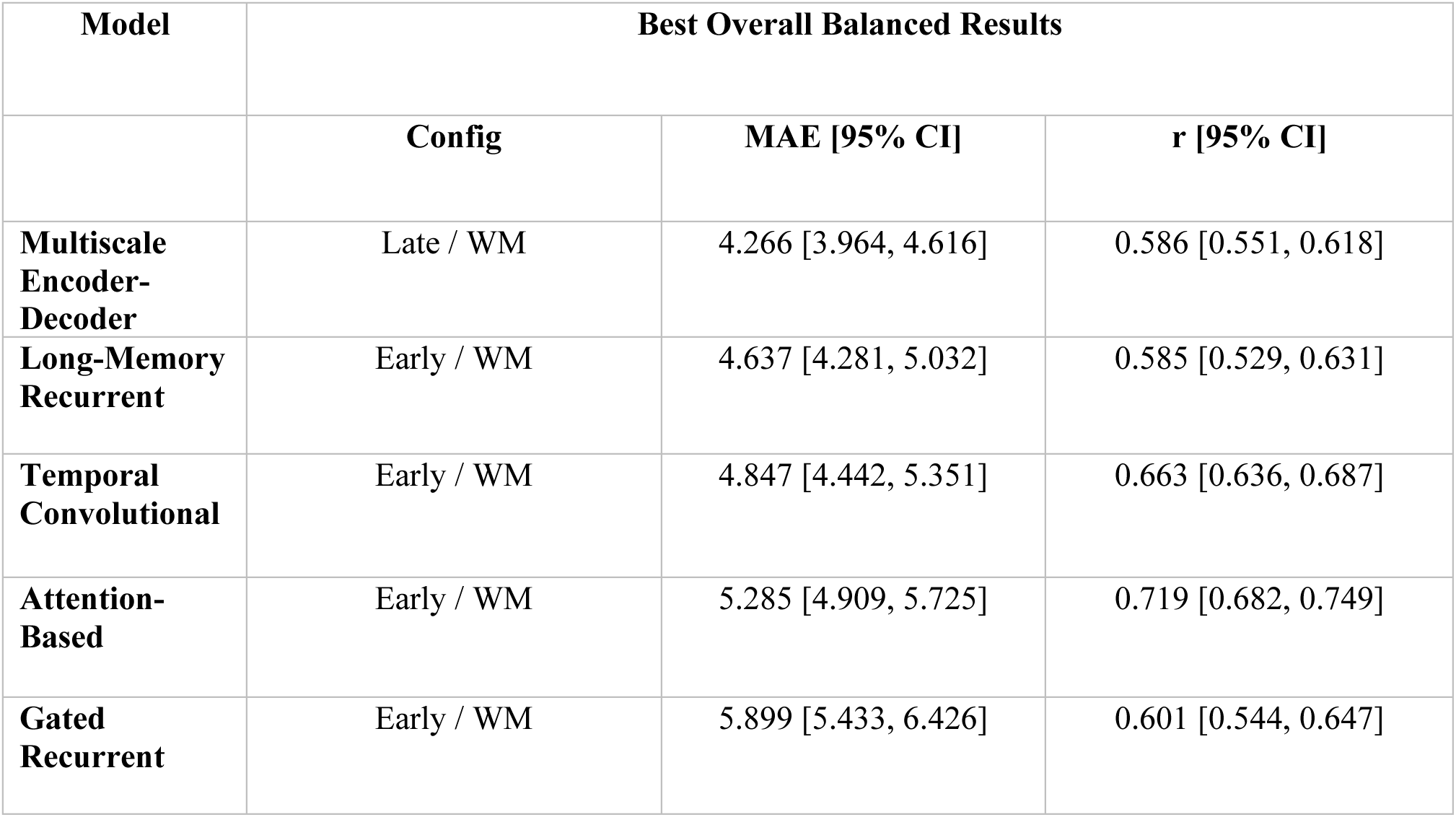
External validation performance. Configuration yielding the optimal balanced result per model, selected using PMAP internal-validation MAE and r only (no MIMIC-III information used in selection). Models are ordered from lowest to highest external MAE; the first three meet the <5 mmHg benchmark. **WM =** waveform-morphology objective; **BRR =** baseline robust regression; **WRR =** weighted robust regression. 95% bootstrap confidence intervals. Clinically motivated benchmark: <5 mmHg MAE.

**Table 3.** External validation performance. Configuration yielding the optimal balanced result per model when selection used the external (MIMIC-III) MAE and r. Because selection used the evaluation data, these results are an optimistic upper bound and are reported for comparison with the internally selected primary results in Table 2. Models are listed in the same order as Table 2. **WM =** waveform-morphology objective; **BRR =** baseline robust regression; **WRR =** weighted robust regression. 95% bootstrap confidence intervals. Clinically motivated benchmark: <5 mmHg MAE.

| **Model** | **Best Overall Balanced Results, Externally Selected (Optimistic Upper Bound)** |  |  |
|---|---|---|---|
|  | **Config** | **MAE [95% CI]** | **r [95% CI]** |
| **Multiscale Encoder-Decoder** | Early / BRR | 4.493 [4.145, 4.879] | 0.721 [0.679, 0.755] |
| **Long-Memory Recurrent** | Late / WM | 4.353 [4.020, 4.702] | 0.656 [0.624, 0.686] |
| **Temporal Convolutional** | Late / WM | 4.358 [4.036, 4.736] | 0.599 [0.563, 0.633] |
| **Attention-Based** | Early / BRR | 4.946 [4.602, 5.390] | 0.722 [0.683, 0.752] |
| **Gated Recurrent** | Late / WM | 4.276 [3.954, 4.633] | 0.619 [0.590, 0.644] |

Differences were also seen across fusion strategies and loss functions. Additional performance metrics and analyses are provided in **Supplementary Table 5.**

The internal-to-external performance gap is seen in Fig. 2. For example, the temporal convolutional model increased from 2.005 to 4.847 mmHg MAE, and the attention-based model increased from 2.302 to 5.285 mmHg. Pearson correlation also decreased for every architecture, from 0.805 to 0.663 (temporal convolutional), 0.761 to 0.586 (multiscale encoder-decoder), 0.695 to 0.585 (long-memory recurrent), and 0.689 to 0.601 (gated recurrent), with the smallest change for the attention-based model (0.758 to 0.719).

**Fig. 2.**
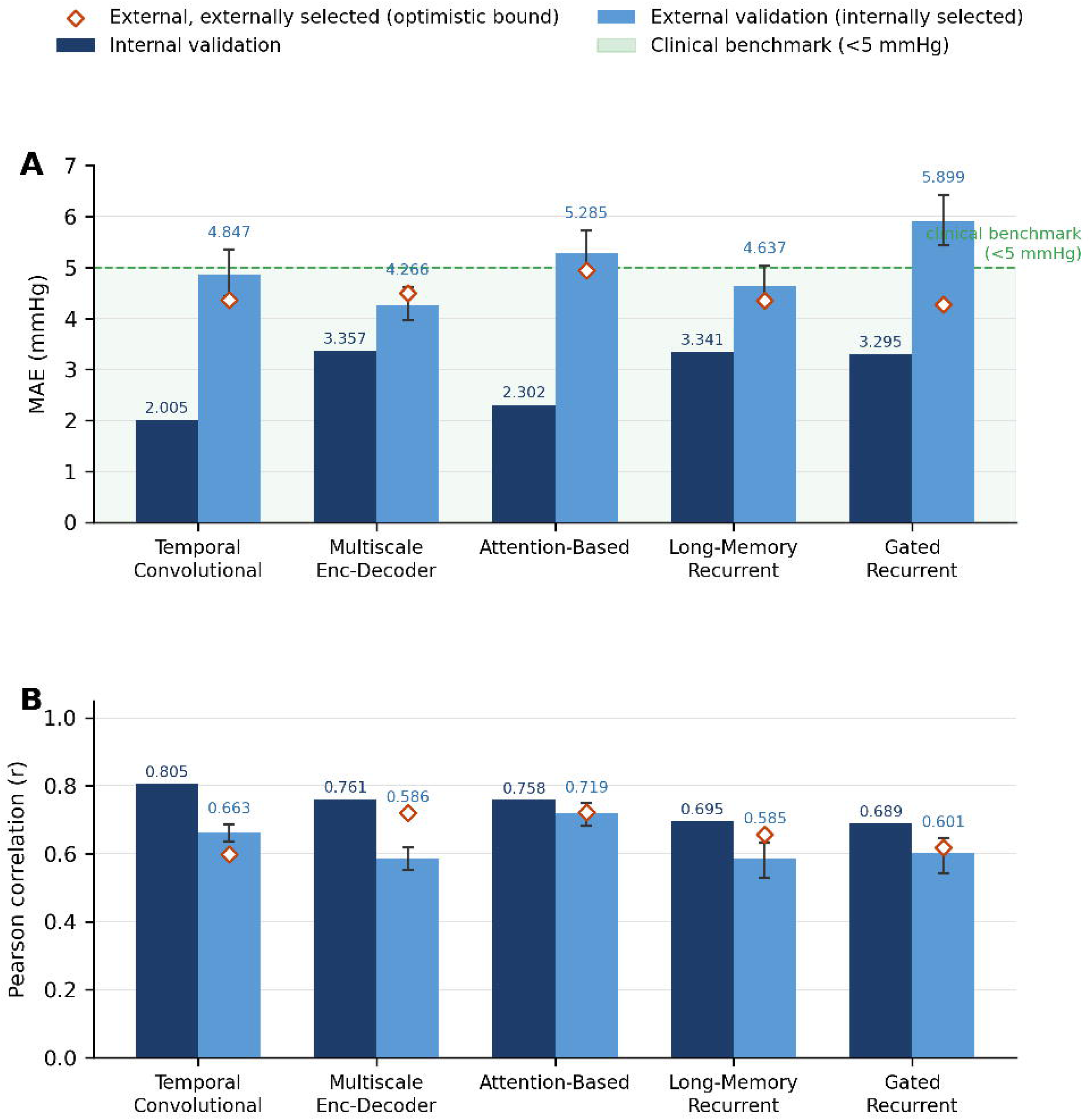
Internal and external validation performance of the balanced model configurations, selected for each architecture using PMAP internal-validation metrics only. a) Mean absolute error (MAE) in mmHg for the balanced models. The clinically motivated benchmark of <5 mmHg is highlighted in green. Internal estimates derive from a segment-level split and are optimistic. Error bars show 95% patient-level bootstrap confidence intervals for external performance; diamonds mark the external performance of the externally selected configurations (Table 3, optimistic upper bound). b) Pearson correlation (r) for the selected configuration of each architecture.

Performance also varied by primary diagnosis category, age group, and sex (see **Supplementary Tables 13–14** for details). Among diagnostic groups, the trauma subgroup (n=5) had the highest MAE point estimate for two of five architectures (temporal convolutional and attention-based) and the lowest Pearson correlation for all five architectures. Given the small trauma subgroup, these findings should be considered exploratory. The two largest diagnostic groups, subarachnoid hemorrhage/aneurysm (MAE 3.881–5.954 mmHg across architectures) and other intracranial hemorrhage (MAE 4.421–5.850 mmHg), produced results that generally tracked the overall cohort estimates. No architecture was uniformly best across the evaluated subgroups, and the age- and sex-stratified results should also be interpreted cautiously given subgroup sizes and overlapping confidence intervals.

Patient-averaged performance (Supplementary Table 15) was modestly worse than the segment-weighted values reported above, with patient-averaged mean MAE exceeding the 5-mmHg benchmark for all five architectures. Meanwhile, the patient-averaged median remained below the benchmark for three of five architectures and closely tracked the segment level mean MAE, indicating a right-skewed distribution of MAE per patient. This divergence was only weakly and inconsistently associated with a patient’s representation of hypertensive-range segments (per-patient MAE of the segment-mean ICP estimate versus the percentage of segments with mean reference ICP >15 mmHg): the association was positive for three of five architectures (Spearman ρ=0.21–0.31, p=0.001–0.024), absent for the long-memory recurrent model (ρ=−0.01, p=0.89), and negative for the gated recurrent model (ρ=−0.29, p=0.002).

### Beat-Level Peak Morphology

To assess whether reconstructed waveforms preserved beat-level morphology, we detected the P1, P2, and P3 peaks in reference and predicted beats from 10,030 external segments across all 111 patients (Supplementary Table 17, Supplementary Fig. 3). P1 and P2 were identifiable in only 42.6% of reference beats. For these beats, the multiscale encoder-decoder and long-memory recurrent models almost never produced a distinguishable P1–P2 pair (3.0% and 1.2%), whereas the temporal convolutional (74.9%), attention-based (41.6%), and gated recurrent (17.1%) models did so more often. Where peaks were detected, agreement with the reference was weak. P2/P1 ratio correlation ranged from 0.14 (temporal convolutional) to 0.40 (gated recurrent), above a shuffled null (0.03–0.12), but P2/P1 ratio and P1 latency errors were larger than those of a constant-value baseline for every model. P1-dominance classification was below the majority-class rate for three models and only marginally above it for the other two, and P3 was not reliably detected (sensitivity ≤0.08 for four of five models; temporal convolutional 0.49 sensitivity with 0.57 specificity). The models therefore captured mean pressure level and the dominant pulsatile component, but not detailed peak configuration.

### Stratified Performance Across the ICP Range

To assess whether external validation performance was uniform across the clinical ICP range, Bland-Altman analysis was performed for each model’s internally selected configuration, stratified by ICP bin. The results for this analysis are found in Fig. 5 and Table 4. Systematic bias in the normotensive range (≤15 mmHg) was small for the temporal convolutional, multiscale encoder-decoder, and attention-based models (−0.5 to +1.4 mmHg), whereas the recurrent models overpredicted (long-memory +3.1 mmHg, gated recurrent +5.3 mmHg). All five models showed substantial underprediction bias above 15 mmHg, consistent with regression toward the normotensive mean under a skewed training distribution. This pattern was consistent across architectures, with biases ranging from −3.0 to −7.2 mmHg in segments with mean reference ICP >15 to ≤20 mmHg, from −8.7 to −13.2 mmHg in segments with mean reference ICP >20 to ≤25 mmHg, and from −15.7 to −20.7 mmHg in segments with mean reference ICP >25 mmHg. The temporal convolutional model exhibited the widest limits of agreement across elevated ICP bins (widths 16.5–18.9 mmHg versus 6.6–15.6 mmHg for the other models). The temporal convolutional and attention-based models had largely mean-independent overall error distributions (heteroscedasticity r=−0.17 and −0.13, respectively), whereas error was strongly mean-dependent for the multiscale encoder-decoder (r=−0.86), long-memory recurrent (r=−0.93), and gated recurrent (r=−0.60) models. These findings indicate that the clinically motivated benchmark met by three of five architectures in **Table 2** reflects average performance across a predominantly normotensive external cohort and does not extend uniformly to the hypertensive range, where clinical decision-making is most consequential.

**Table 4.** Stratified Bland-Altman analysis by ICP bin for each model. Bias and limits of agreement (LoA) are reported for normotensive (≤15 mmHg), mildly elevated (15–20 mmHg), moderately elevated (20–25 mmHg), and severely elevated (>25 mmHg) ranges. n=606,280 segments.

| Model | ICP Bin | n segments | Bias | LoA Lower | LoA Upper |
|---|---|---|---|---|---|
| **Multiscale Encoder-Decoder** | **≤ 15 mmHg** | **538,591** | 1.414 | -5.631 | 8.459 |
|  | **>15 to ≤20 mmHg** | **49,275** | -6.944 | -10.687 | -3.201 |
|  | **>20 to ≤25 mmHg** | **13,668** | -12.261 | -15.906 | -8.616 |
|  | **> 25 mmHg** | **4,746** | -18.977 | -24.414 | -13.541 |
| **Long-Memory Recurrent** | **≤ 15 mmHg** | **538,591** | 3.077 | -3.973 | 10.127 |
|  | **>15 to ≤20 mmHg** | **49,275** | -5.545 | -8.86 | -2.23 |
|  | **>20 to ≤25 mmHg** | **13,668** | -10.85 | -14.238 | -7.461 |
|  | **> 25 mmHg** | **4,746** | -17.696 | -23.123 | -12.269 |
| **Temporal Convolutional** | **≤ 15 mmHg** | **538,591** | -0.479 | -9.93 | 8.972 |
|  | **>15 to ≤20 mmHg** | **49,275** | -7.168 | -15.408 | 1.072 |
|  | **>20 to ≤25 mmHg** | **13,668** | -13.227 | -22.68 | -3.773 |
|  | **> 25 mmHg** | **4,746** | -20.689 | -29.333 | -12.044 |
| **Attention-Based** | **≤ 15 mmHg** | **538,591** | 1.074 | -9.677 | 11.825 |
|  | **>15 to ≤20 mmHg** | **49,275** | -7.165 | -14.988 | 0.657 |
|  | **>20 to ≤25 mmHg** | **13,668** | -13.121 | -19.858 | -6.383 |
|  | **> 25 mmHg** | **4,746** | -19.154 | -26.547 | -11.762 |
| **Gated Recurrent** | **≤ 15 mmHg** | **538,591** | 5.314 | -2.626 | 13.254 |
|  | **>15 to ≤20 mmHg** | **49,275** | -2.975 | -8.383 | 2.434 |
|  | **>20 to ≤25 mmHg** | **13,668** | -8.743 | -14.924 | -2.562 |
|  | **> 25 mmHg** | **4,746** | -15.726 | -22.356 | -9.097 |

## Discussion

This study demonstrates that deep learning models trained on routinely collected physiological waveform data can not only estimate scalar ICP but also reconstruct continuous ICP waveforms with MAE below the clinically motivated accuracy threshold of 5 mmHg [18, 19] for three of five architectures when configurations were selected using internal validation data only (4.266–4.847 mmHg), while Pearson correlation (r = 0.585–0.719) reflects a moderate degree of waveform-level temporal agreement with reference signals, although beat-level peak morphology (P1/P2/P3) was not reliably recovered (Supplementary Table 17). Prior work [14, 15, 16] has demonstrated single-institution ICP waveform reconstruction. We extend this work with cross-institutional external validation using only routinely acquired bedside signals across 111 held-out patients. Across the five tested model architectures, three (the multiscale encoder-decoder, long-memory recurrent, and temporal convolutional models) had point-estimate external validation MAEs below the 5-mmHg benchmark when configurations were selected using internal data only, whereas the attention-based (5.285 mmHg) and gated recurrent (5.899 mmHg) models did not. Under external selection (Table 3, optimistic upper bound), all five were below the benchmark (4.276–4.946 mmHg), which is within the accuracy range of invasive intraparenchymal monitors. These external validation results exceed our group’s prior internally validated work (Nair et al. [16]), which also evaluated performance across a full patient cohort using extracranial signals, in Pearson correlation for all five architectures (r = 0.585–0.719 versus 0.55–0.58) and in MAE for three of five (the attention-based and gated recurrent MAEs were higher than the prior 4.98–5.20 mmHg internal MAEs). Other recent studies in this space, like Lei et al. [14] (ABP-only) and Shu et al. [17] (ECG+ABP fusion), report lower segment-level MAE, but via single-patient or small-cohort demonstrations without cohort-level external validation. Table 5 further highlights the larger scale of the present evaluation: prior ICP waveform-reconstruction studies included cohorts of 10–11 patients, whereas the present study included 47 development patients and 111 external-validation patients (158 patients total). Cross-study MAE and Pearson correlation values should be interpreted cautiously because metric definitions and aggregation procedures differ. None of these studies, including our own prior work, tested whether a trained model generalizes to an independent institution’s data, a gap that this study begins to address.

**Table 5.** Comparison with prior ICP waveform-reconstruction studies. Cohort size reports analyzed patients. MAE values are in mmHg. Internal validation denotes evaluation within the study dataset; external validation denotes evaluation on an independent cohort or dataset. Reported MAE and Pearson r values follow the definitions and aggregation procedures used in each study and should therefore be interpreted cautiously when compared across studies. The present study includes 47 development patients and 111 external-validation patients; metric ranges span the five internally selected configurations. NR indicates that the metric was not reported in the original study. Dashes indicate that external-validation metrics were not applicable because external validation was not performed. TCD-CBFV = transcranial Doppler–derived cerebral blood flow velocity.

| Study | Cohort, n | Inputs | External Validation | Internal MAE | Internal r | External MAE | External r |
|---|---|---|---|---|---|---|---|
| **Lei et al. [14]** | 10 | ABP | No | 0.42 ± 0.18 | 0.822 | — | — |
| **Megjhani et al. [15]** | 11 | ABP + ECG + TCD-CBFV | No | 3.88–3.94 | NR | — | — |
| **Nair et al. [16]** | 10 | ABP + PPG + ECG | No | 4.98–5.20 | 0.55–0.58 | — | — |
| **Shu et al. [17]** | 10 | ECG + ABP | No | 0.359 | 0.932 | — | — |
| **Present study** | **158 (47 + 111)** | **ABP + PPG + ECG** | **Yes** | **2.005–3.357** | **0.689–0.805** | **4.266–5.899** | **0.585–0.719** |

The multiscale encoder-decoder achieved the lowest external MAE (4.266 mmHg), followed by the long-memory recurrent (4.637 mmHg) and temporal convolutional (4.847 mmHg) architectures, whereas the attention-based architecture achieved the highest external Pearson r (0.719) at a higher MAE (5.285 mmHg). No architecture led on both metrics, as the internally selected multiscale encoder-decoder configuration had a correlation of 0.586, whereas its externally selected configuration reached r = 0.721 (Table 3). Both dimensions are clinically meaningful, as downstream utilities including pulse amplitude computation and B-wave detection depend on waveform shape as well as pressure accuracy. Because peak-level morphology was not recovered (see limitations below), these downstream uses remain untested. The observation that MAE and Pearson r do not consistently align across architectures reinforces that neither metric alone is sufficient to characterize reconstruction quality, and both should be reported as complementary evaluation criteria. Late fusion tended to yield lower external MAE (significantly so for the temporal convolutional and long-memory recurrent architectures, and for the gated recurrent architecture under two of three objectives, in patient-level paired testing, Supplementary Table 7), whereas early fusion more often preserved waveform correlation; this pattern was not uniform across architectures (no significant difference for the multiscale encoder-decoder or attention-based models), and may reflect how each strategy handles cross-channel information under distributional shift. Across objective functions, the waveform-morphology objective appeared most consistently among the lowest-MAE and balanced configurations. In contrast, the baseline robust regression objective more often characterized the highest-correlation settings, each consistent with its design intent, and the weighted robust regression objective did not emerge as the top configuration in any panel. These objective-level patterns are descriptive: after correction for multiple comparisons, patient-level pairwise differences between objectives were significant for only a minority of comparisons (Supplementary Table 8). Selecting configurations on the external data, as in Table 3, lowered external MAE by 0.3–1.6 mmHg for four of five architectures (the multiscale encoder-decoder was the exception, at 0.2 mmHg higher), illustrating the optimism of selecting on the evaluation data and why internally selected configurations are reported as the primary results. While all models exhibited an internal-to-external performance gap, this is expected under genuine cross-institutional domain shift. Moreover, the fact that three of five models met the clinically motivated benchmark on this largely normotensive external dataset despite this performance gap, without any external information used in model selection, is consistent with the possibility that the learned relationships they rely on can generalize across institutions, at least for some architectures.

We regard the multiscale encoder-decoder, long-memory recurrent, and temporal convolutional models as the principal architectures because they were the three that met the benchmark when configurations were selected on internal data only. A plausible, though untested, explanation is that their inductive biases suit the structure of ICP waveform reconstruction. ICP is a quasi-periodic signal whose pulse morphology is shaped by cardiac-cycle dynamics over multiple time scales. Multiscale encoder-decoder and dilated temporal convolutional architectures capture such structure through hierarchical or dilated receptive fields, and a long-memory recurrent network can carry information across the full segment. The gated recurrent model has a shorter effective memory and was the most configuration-sensitive (external MAE 4.28–5.90 mmHg across fusion and objective configurations), while the attention-based model has weaker built-in locality and ordering priors and may need more training data than our 47-patient development cohort, although it achieved the highest correlation. These explanations are merely hypotheses, however, as we did not perform ablations that isolate individual architectural properties.

Representative waveforms illustrate these findings (Figs. 3 and 4). For a pathologically altered, P2-dominant segment (Fig. 3), the gated recurrent model followed pulse amplitude and timing most closely but merged P1, P2 and P3 into a single rounded peak. The multiscale encoder-decoder and attention-based models produced a single smoothed pulse of reduced amplitude. The temporal convolutional model followed the pulse with high-frequency noise, and the long-memory recurrent model returned a nearly flat trace. For a P1-dominant segment with higher mean ICP (Fig. 4), all models underestimated the mean level by approximately 2–4 mmHg, and four of five reproduced only small-amplitude pulses; the gated recurrent model recovered pulse amplitude, but its peaks were slightly delayed relative to the reference. These are single segments chosen for illustration and should not be taken as representative of all segments; the beat-level analysis in Supplementary Table 17 provides the quantitative assessment.

**Fig. 3.**
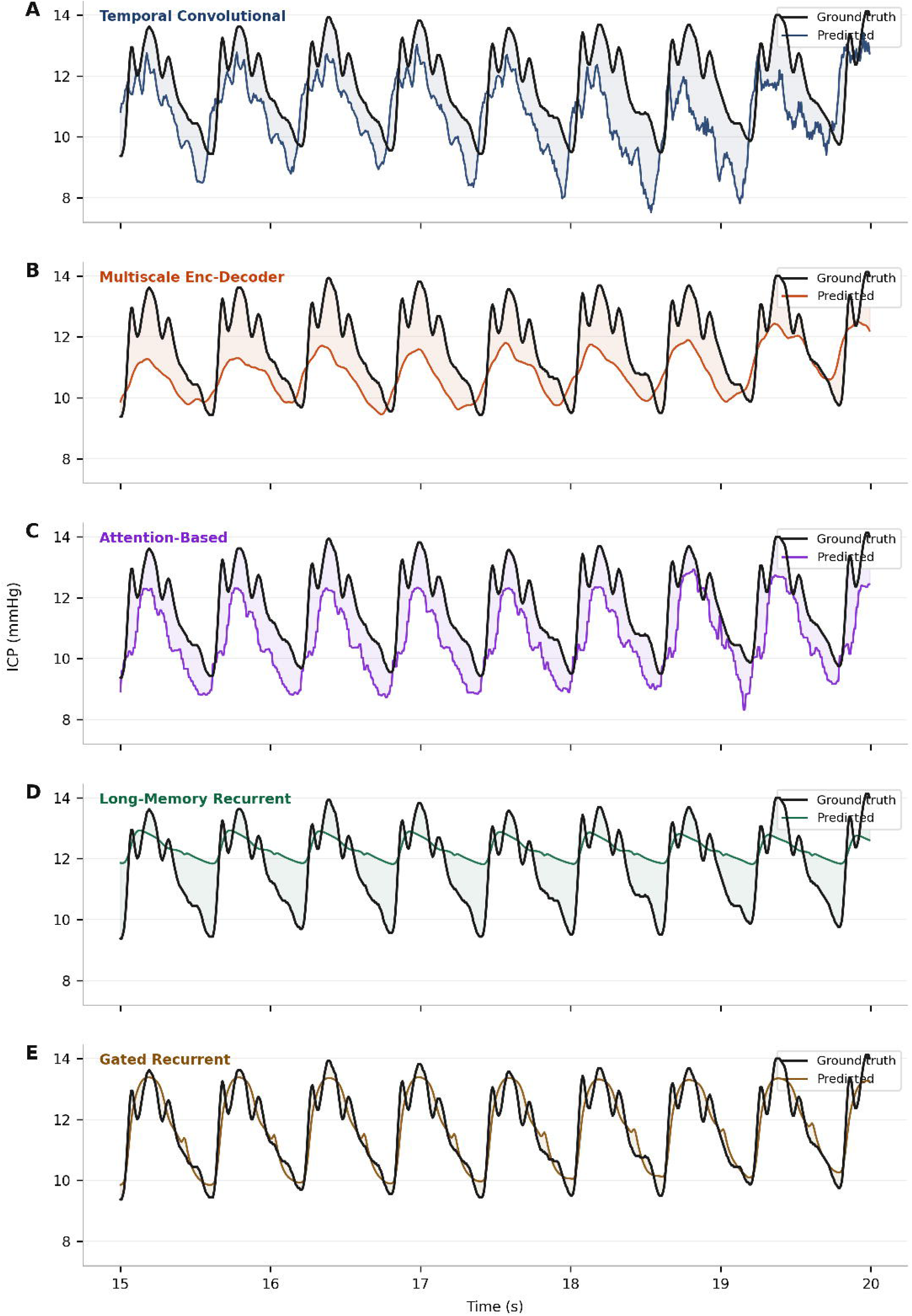
Representative ICP waveform predictions on a pathologically altered ground truth waveform (representative segment from the external validation cohort), from the internally selected balanced a) Temporal Convolutional Model, b) Multiscale Encoder-Decoder Model, c) Attention-Based Model, d) Long-Memory Recurrent Model, and e) Gated Recurrent Model.

**Fig. 4.**
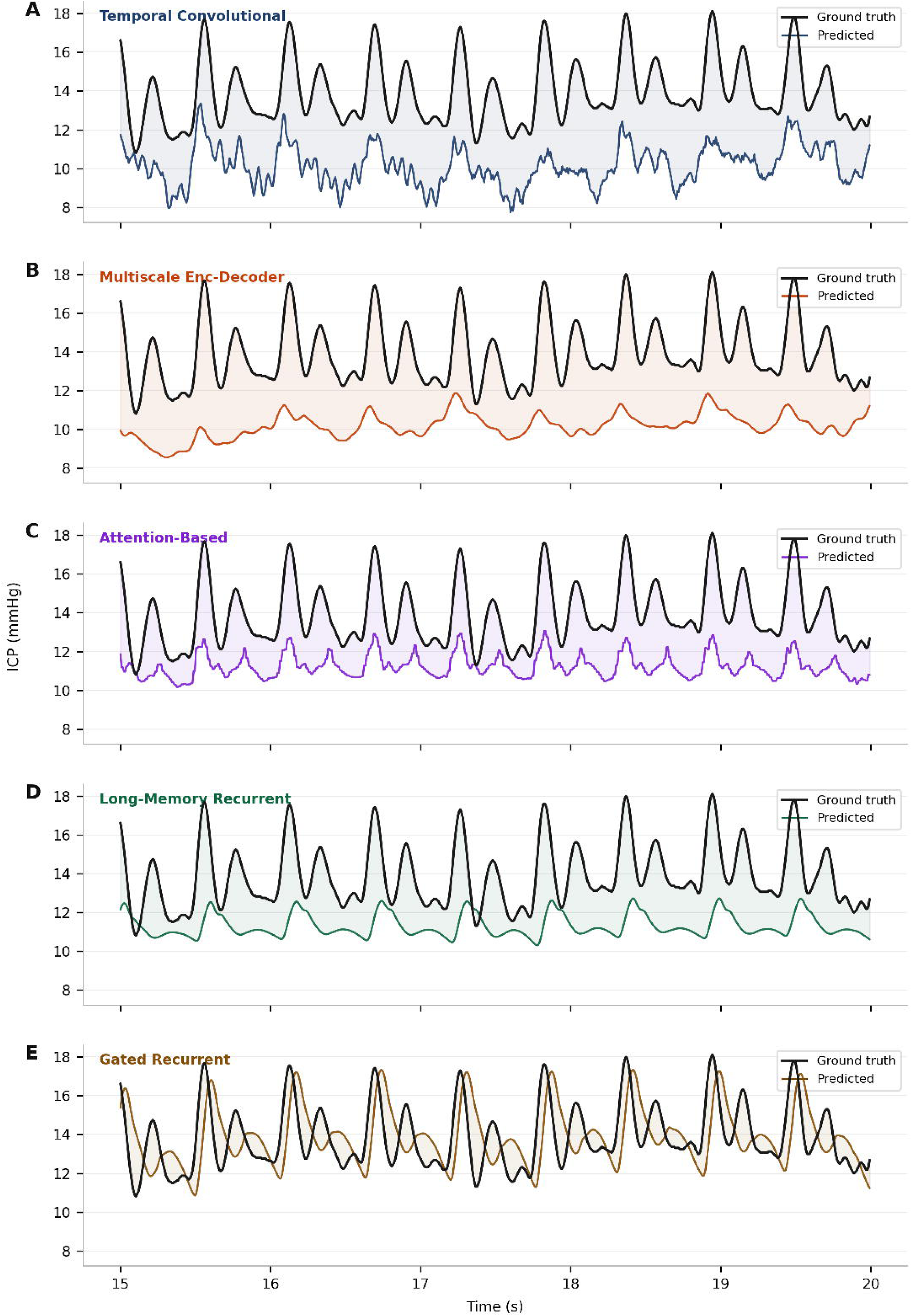
Representative ICP waveform predictions on a normal (P1-dominant) ground truth waveform (representative 5-second segment from the external validation cohort), from the internally selected balanced a) Temporal Convolutional Model, b) Multiscale Encoder-Decoder Model, c) Attention-Based Model, d) Long-Memory Recurrent Model, and e) Gated Recurrent Model.

The present findings have implications for ICU applicability and access to monitoring. A key strength of this framework is that it uses only ABP, PPG, and ECG, which are routinely acquired in intensive care, without requiring operator-dependent measurements, specialized imaging, or new bedside hardware. This strength makes the approach more practical than many alternative ICP-estimation strategies and easier to integrate into existing monitoring workflows. The fact that the three principal architectures (multiscale encoder-decoder, long-memory recurrent, and temporal convolutional) met the benchmark under external validation without any external information used in selection suggests that this framework may be robust enough to serve as an adjunctive tool across heterogeneous care environments. With additional validation, this approach could provide basic physiologic context when invasive ICP monitoring is unavailable or infeasible, potentially supporting earlier recognition of worsening intracranial dynamics, longitudinal trending, and more informed triage for escalation of care.

However, the largest limitation of this study is that performance was not uniformly robust across all ICP ranges. As evidenced by the Bland-Altman analysis, while the temporal convolutional, multiscale encoder-decoder, and attention-based models showed minimal bias in the normotensive range, every model showed increased underprediction with rising ICP. This limits the direct applicability of our models specifically in hypertensive ranges, where clinical decision-making is most consequential.

The cause for this limitation is likely the significantly lower representation of hypertensive segments. In our MIMIC-III cohort, only 11.2% of all segments had mean ICP above 15 mmHg, and segments with significant hypertension (>25 mmHg) comprised less than 1% of our data (see exact counts in **Table 4**). This skew reflects ICU epidemiology, as intracranial hypertension is relatively rare among monitored patients and episodes are generally treated promptly. Prior studies have navigated this concern in different ways. Our own group’s earlier study [16] enriched training data by enforcing a minimum duration of hypertensive recording, improving the representation of hypertensive patients in the dataset. However, this came at the cost of reducing cohort size from 116 to 19 patients, of whom 10 were analyzed in the MIMIC-III Waveform Matched dataset. Since this sample size is incompatible with a multi-patient external validation study, we chose to eliminate this restriction. Lei et al. [14] also reported degraded accuracy at elevated ICP, with classification accuracy dropping to 84.3% with an underestimation rate of 15.7% at the highest ICP ranges above 40 mmHg. The authors similarly noted that larger datasets are needed to improve reliability in this regime. The present study adds stratified reporting with multi-institutional external validation in this space. This limitation is shared by prior work [14, 16] and will require analysis of hypertension-enriched multi-centre datasets to overcome.

A second limitation is that the models did not reproduce beat-level pulse morphology. In an analysis of the canonical cardiac peaks (Supplementary Table 17), the multiscale encoder-decoder and long-memory recurrent models almost never produced a distinguishable P1–P2 structure, and for the other models agreement with reference peak amplitude ratios and latencies was weak and did not improve on a constant-value baseline; P3 was not reliably detected. The correlation values reported here therefore appear to reflect agreement in the dominant cardiac-frequency oscillation rather than detailed peak configuration, and applications that depend on pulse morphology, such as P2/P1-based assessment of intracranial compliance, are not supported by the present models. Because inter-signal timing was not corrected, latency errors may be partly attributable to timing offsets, although amplitude-ratio errors would not be.

Additional limitations bound these conclusions. First, the segment-level train/validation split allowed segments from the same patient to contribute to both partitions and will therefore yield optimistic internal validation performance relative to a fully patient-independent split. A patient-level split was not used because the limited PMAP cohort size (n = 47) would have produced small validation partitions with estimates highly sensitive to individual patient assignment. Importantly, this limitation applies to the internal validation only; the MIMIC-III cohort was entirely independent and serves as the primary assessment of cross-patient and cross-institutional generalization. Second, PMAP’s low segment survival rate after quality control (6.9%) reflects the noise of real-world bedside recordings relative to the comparatively curated MIMIC-III release, and performance in noisier deployment environments may differ accordingly. Additionally, the PMAP development cohort was derived from a convenience sample of patients with simultaneous high-resolution ICP, ABP, PPG, and ECG recordings. As a result, the cohort may not fully represent the broader neurological and neurosurgical patient population, particularly given differences in the clinical indications for invasive ICP monitoring and the availability of high-resolution PPG recordings. The proportion of patients within the relevant clinical population who met these recording requirements could not be determined from the available data. Third, a fully non-invasive ICP reconstruction paradigm would require modality-specific ablation (i.e. exclusion of ABP) under pairwise signal combinations, which we plan to address in a future analysis. Fourth, physiologic inter-signal timing delays (for example, ECG-to-pressure transit times) were not corrected and were retained to preserve native timing relationships between signals. Because such delays may differ across institutions and monitoring systems, uncorrected timing differences may contribute to the internal-to-external performance gap, particularly for correlation-based metrics. Finally, the external validation cohort was derived from MIMIC-III recordings acquired between 2001 and 2012, whereas the PMAP training data were recorded between 2016 and 2021. Differences in monitoring hardware, ICU practices, and patient populations across these eras may therefore contribute to domain shift and may limit direct generalizability to contemporary clinical environments. Specific clinical management guidelines in effect at each institution during the respective recording periods were not documented and could not be directly compared. However, given the temporal and institutional separation, some divergence in practice patterns is plausible. At the same time, the use of more contemporary PMAP data for model development and successful validation on an older, independent cohort suggests that the learned relationships were not specific to a single acquisition era or, by extension, a single set of institutional practice patterns. Further validation in contemporary external cohorts will be needed to confirm performance under current monitoring and care practice. Despite the listed limitations, this study represents a meaningful step forward, demonstrating that models trained at one institution can generalize to an independent institution’s data, a precondition for robust and standardized extracranial ICP estimation. Building on this foundation, future work incorporating larger and more hypertension-representative cohorts will be essential to extend these findings toward a clinically deployable tool across the full spectrum of intracranial pressure.

## Methods

### Data Sources and Inclusion Criteria

Models were trained via post hoc analysis of a convenience sample of continuous physiologic data from PMAP at Johns Hopkins Hospital. Patients were included if they had simultaneous and continuous recordings of ICP, invasive ABP, PPG, and ECG. After preprocessing and quality filtering, 47 patients remained across 91 recording sessions, yielding approximately 270 hours of data comprising 32,446 thirty-second segments. PMAP waveform data were recorded between 2016 and 2021. Bedside physiologic signals were acquired using GE HealthCare CARESCAPE multiparameter monitors (GE HealthCare, Chicago, IL, USA). High-resolution waveforms and numerics were captured and time-aligned using the Sickbay clinical platform (Medical Informatics Corp., Houston, TX, USA). Detailed characterization of all recording equipment was not available. Use of this dataset was approved by the Johns Hopkins Medicine Institutional Review Board (IRB; protocol No. IRB00320184).

Model external validation was accomplished using data from the MIMIC-III Waveform Database acquired at Beth Israel Deaconess Medical Center in Boston, Massachusetts, drawing from the MIMIC-III Waveform Matched Subset. After preprocessing and quality filtering, 111 patients remained across 222 recording sessions, yielding approximately 5,052 hours of data comprising 606,280 thirty-second segments. All data from the MIMIC-III Waveform Database was recorded between 2001 and 2012.

Because MIMIC-III is a publicly available, de-identified database, use of these data was exempt from additional IRB review. **Fig. 1** summarizes the data sources, preprocessing pipeline, model configurations, and validation strategy. Segment counts per patient were right-skewed for both cohorts (Supplementary Figs. 1–2); a sensitivity analysis excluding MIMIC-III patients with fewer than 100 segments showed negligible effect on external validation performance (max ΔMAE = 0.0009 mmHg, max Δr = 0.0002). Patient-level bootstrapping was used throughout to account for within-patient segment correlation when estimating confidence intervals; however, because segment-weighted point estimates still reflect greater contribution from patients with longer recordings, we additionally report patient-averaged performance in the Results.

### Data Preprocessing and Quality Control

An identical preprocessing pipeline was applied to both datasets. Native sampling rates differed between datasets. MIMIC-III signals were sampled at 125 Hz for all four channels. PMAP signals were originally sampled at 240 Hz (ECG), 120 Hz (ICP and ABP), and 60 Hz (PPG), and were provided already upsampled to 240 Hz, the highest native rate. All recordings were resampled to a common 125 Hz, the lower of the two datasets’ rates. In both datasets the four signals were provided temporally aligned, sharing a common time index with each sample recorded simultaneously. No correction was applied for physiologic inter-signal delays (for example, ECG-to-pressure transit times), so as to preserve the native timing relationships between signals. Continuous recordings were then segmented into non-overlapping 30-second segments (3,750 samples). Entire segments were rejected on any quality failure, rather than excising individual samples. Quality control proceeded in two passes. Pass 1 conducted physiologic checks, and rejected segments if they contained any NaN values, if ICP samples fell outside [−10, 40] mmHg or ABP samples outside [20, 300] mmHg, if any channel exhibited insufficient peak-to-peak amplitude variation across 2-second sub-windows (indicating flat or stuck signals), if the ICP signal lacked cardiac periodicity as assessed by Welch power spectral density (requiring either a minimum band signal-to-noise ratio (SNR) of 3 dB or at least 15% of total power in the 0.8–2.5 Hz cardiac band), if ECG-derived heart rate via GQRS R-peak detection fell outside [10, 250] bpm, or if the broadband-to-dominant-frequency spectral power ratio exceeded 4.25 (indicating excessive noise). Pass 2 applied ICP-specific morphology filters to segments surviving Pass 1: segments were rejected if the ICP channel contained fewer than 100 distinct amplitude values (targeting quantized or stuck-signal artifacts), or if the ratio of noise peaks to physiologic peaks exceeded 4.00, where physiologic peaks were defined by prominence ≥ 1.0 and minimum spacing of 50 samples, and noise peaks by prominence ≥ 0.05 and minimum spacing of 2 samples. No signal filtering was applied beyond these quality-control criteria.

Of the 467,699 raw PMAP segments, 32,446 (6.9%) survived both passes, compared to 1,071,698 of 2,904,410 raw MIMIC-III segments (36.9%) of which 606,280 (20.9%) belonged to patients with linked clinical records and constitute the external validation set; this reflects the substantially less curated nature of real-world bedside recordings relative to the curated MIMIC-III release. Per-filter rejection statistics for both datasets are provided in **Supplementary Tables 1 and 2**, and a diagram of patient level flow is presented in Fig. 6.

The two dominant rejection sources in PMAP were pre-resampling NaN values (38.1%) and the ICP-specific noise-to-physiologic-peak ratio filter (40.3%). Per-channel analysis showed NaN-driven rejection was dominated by the ABP channel in PMAP (91.7% of NaN-affected segments) but by the ICP channel in MIMIC-III (13.4%), with correspondingly different gap structures: PMAP’s ABP dropouts were near-uniformly full-segment, consistent with structural recording gaps rather than brief signal loss, while MIMIC-III’s NaN gaps were substantially shorter (median 0.21 s), making short window imputation inapplicable to PMAP but a plausible target for future preprocessing refinement in the MIMIC-III cohort specifically (**Supplementary Tables 3–4**). The ICP-specific noise-to-physiologic-peak ratio filter is worth noting in the context of clinical deployment: since the model is designed to serve as an alternative to invasive ICP monitoring, the invasive ICP signal would not be required as a model input in practice, which would eliminate this dominant source of data attrition. For deployment, quality control would instead need to focus on the integrity of the extracranial input signals (ABP, PPG, and ECG). These signals underwent fewer signal-specific quality checks than ICP in the present preprocessing pipeline; therefore, their lower rejection rates should not be interpreted as evidence of inherently greater signal quality.

### Train / Validation / Test Split

PMAP segments, defined as non-overlapping 30-second waveform windows, were divided into 13 equal parts at the segment level, with 10 allocated to training and 3 to internal validation (77/23 split). All MIMIC-III data were held out exclusively for external validation. A segment-level split was used because the limited PMAP cohort size (n = 47) would result in small patient-level training and validation partitions, with performance estimates potentially sensitive to the particular patients assigned to each partition. We therefore prioritized the larger segment-level validation set for model development, while reserving the entirely independent MIMIC-III cohort for patient-independent external validation.

### Input Representation and Modality Strategies

Two fusion strategies were evaluated. In early fusion, all three input channels are concatenated and jointly processed by a single shared model. In late fusion, three model instances are trained independently on single-channel inputs and their predictions are averaged at inference.

### Model Training and Evaluation

Sequence-to-sequence models including convolutional, recurrent, and attention-based paradigms were trained under a consistent optimization framework. Models were optimized using three regression-based objective formulations, including a baseline robust objective, a weighted robust objective, and a waveform-morphology objective. Principal architectural details for each of the five model classes, including layer structure and key model dimensions, are provided in **Supplementary Table 16**. The five model architecture classes were selected based on our group’s previous work by Nair et al. [16], which evaluated these approaches for ICP waveform reconstruction. The architectural configurations were subsequently refined through preliminary experiments, during which model hyperparameters, including layer configurations and model dimensions, were iteratively adjusted based on observed internal validation reconstruction performance. Rather than conducting an exhaustive grid search, we used this empirical tuning approach to arrive at the final architectural configurations described in Supplementary Table 16.

Because both datasets were dominated by normotensive ICP values, three regression objectives were compared to examine pointwise amplitude error, elevated-ICP weighting, and waveform morphology. Their mathematical forms are provided below.

For one segment of T samples, let yₜ denote reference ICP and pₜ (equivalently ŷₜ) predicted ICP at time t. Let μᵧ and μₚ denote their respective within-segment means. The objectives below are defined for one segment and averaged across the training batch.

Baseline robust regression (BRR) minimizes the unweighted pointwise log-cosh error:

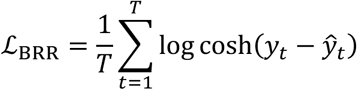

Weighted robust regression (WRR) applies a segment-dependent weight w(μᵧ) to the same pointwise error, increasing the contribution of elevated-ICP segments:

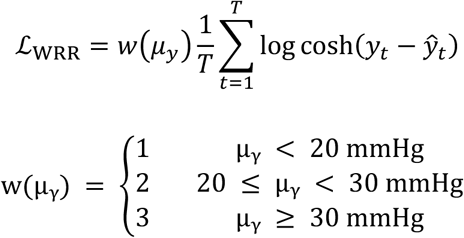

The weight is determined from the mean reference ICP of each 30-second segment (in mmHg) and is applied uniformly to all time points within that segment.

The waveform-morphology (WM) objective comprises a mean-centered waveform-shape term (Lₛ) and a mean-pressure term (Lₘ), combined with the following weighting:

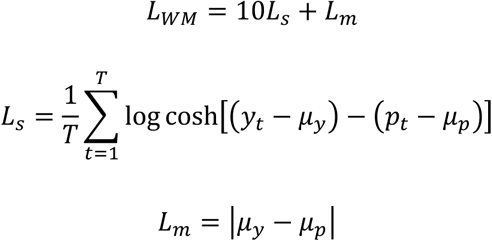

Here, Lₛ measures the log-cosh discrepancy between the reference and predicted waveforms after subtracting their respective segment means, while Lₘ measures the absolute difference in mean ICP. The shape and mean-pressure terms are weighted by 10 and 1, respectively. Unlike the WRR objective, this formulation does not explicitly assign additional weight to elevated-ICP segments. The three objectives were compared to assess how different optimization priorities affect reconstruction under the imbalanced ICP distribution.

Performance was assessed using MAE and Pearson r, computed per segment and averaged. Mean absolute error for a segment of length T was defined as:

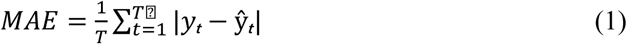

where yₜ and ŷₜ denote the reference and predicted ICP values at time point t, respectively, and T denotes the number of samples in the segment. Pearson correlation for each segment was defined as:

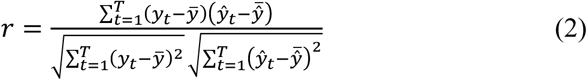

where ȳ and ŷ̄ denote the mean reference and predicted ICP values within the segment, respectively.

For the headline results, we chose to omit any input signal normalization. However, to confirm that this was not a performance-limiting choice, we conducted a normalization ablation comparing z-scored versus unnormalized inputs across the five externally selected configurations (Table 3; not repeated for the internally selected configurations) (**Supplementary Table 9**). To assess whether reported correlations were affected by small temporal misalignment, and to characterize waveform-shape agreement independent of point-error metrics, we additionally computed lag adjusted cross-correlation and dynamic time warping (DTW) distance for all five architectures on the full external validation set (Supplementary Tables 11–12). Beat-level morphology was assessed by detecting the P1, P2, and P3 peaks in reference and predicted beats and comparing peak amplitude ratios, latencies, and intervals against constant-value and shuffled-pair controls (Supplementary Table 17).

### Selection of Best Overall Balanced Result

Best overall balanced results were identified separately for each model architecture by jointly considering internal-validation (PMAP) mean absolute error (MAE) and internal-validation Pearson correlation (r) across all retained early- and late-fusion configurations, so that no external (MIMIC-III) data informed model selection. The MAE penalty, Pearson correlation penalty, and balanced score were computed as follows:

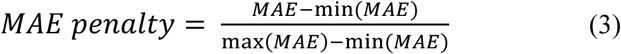

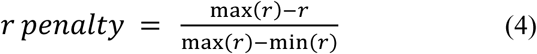

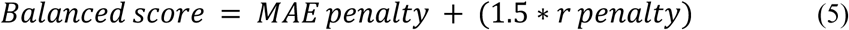

The Pearson penalty was given a higher weight of 1.5 to account for the significance of waveform morphology in our approach. However, to more comprehensively assess sensitivity of model selection to this weighting choice, we repeated configuration selection across a range of weights (1.0×–3.0×; **Supplementary Table 10**). Selection on internal metrics was stable across this full range for four of five architectures; for the multiscale encoder-decoder, the lowest weight (1.0×) favored a different configuration. To quantify the optimism introduced by selecting on the evaluation data, the same procedure was also applied to the external MAE and r, and the resulting externally selected configurations are reported separately as an optimistic upper bound (Table 3).

### Interpretation of Results

Results are interpreted against a clinically motivated benchmark MAE of <5 mmHg, anchored to the in-vivo agreement between invasive intraparenchymal sensors and reference ventricular catheters rather than to their bench specifications [18, 19]. A systematic review and meta-analysis of paired invasive ICP comparisons found a pooled standard deviation of differences of 4.4 mmHg [18], and a single-center observational study and literature review across 76 sources reported an overall in-vivo measurement accuracy of ±6.0 mmHg for intraparenchymal transducers [19]. We note that no formal consensus threshold for non-invasive ICP estimation accuracy has been established. We therefore used an MAE benchmark of <5 mmHg, motivated by reported in-vivo agreement between intraparenchymal ICP sensors and reference ventricular measurements [18, 19]. We also acknowledge that this benchmark is generally useful for a largely normotensive cohort and does not hold for hypertensive regimes. Bench testing of seven transducer types showed errors up to 10 mmHg at high pressures for some devices [20], and a study of one particular monitor type (fiberoptic monitors) in a refractory intracranial hypertension cohort reported substantially larger inter-device discrepancies than general neurocritical care populations (mean 9.2 mmHg, episodes up to 40 mmHg) [21]. However, since our cohort consists largely of normotensive data, we consider a benchmark MAE of <5 mmHg reasonable for our overall results, with the explicit note that it does not apply to hypertensive data.

Uncertainty was quantified via 95% bootstrap confidence intervals (10,000 iterations), computed by resampling patients with replacement to account for within-patient segment correlation. Pairwise model comparisons used the Wilcoxon signed-rank test on per-patient mean MAE differences, with Bonferroni correction for multiple comparisons applied to the model and objective comparisons (see **Supplementary Tables 6 and 8**); the early-versus late-fusion comparisons (Supplementary Table 7) were not corrected and are exploratory.

To assess systematic prediction bias across the clinical ICP range, Bland-Altman analysis was performed for the internally selected configuration of each model on the matched external validation cohort. Segments were assigned to ICP bins according to the mean reference ICP over each 30-second segment: mean ICP ≤15 mmHg, 15 < mean ICP ≤20 mmHg, 20 < mean ICP ≤25 mmHg, and mean ICP >25 mmHg. Bias and 95% limits of agreement were computed overall and within each bin. Results are reported in **Table 4** and **Fig. 5**.

**Fig. 5.**
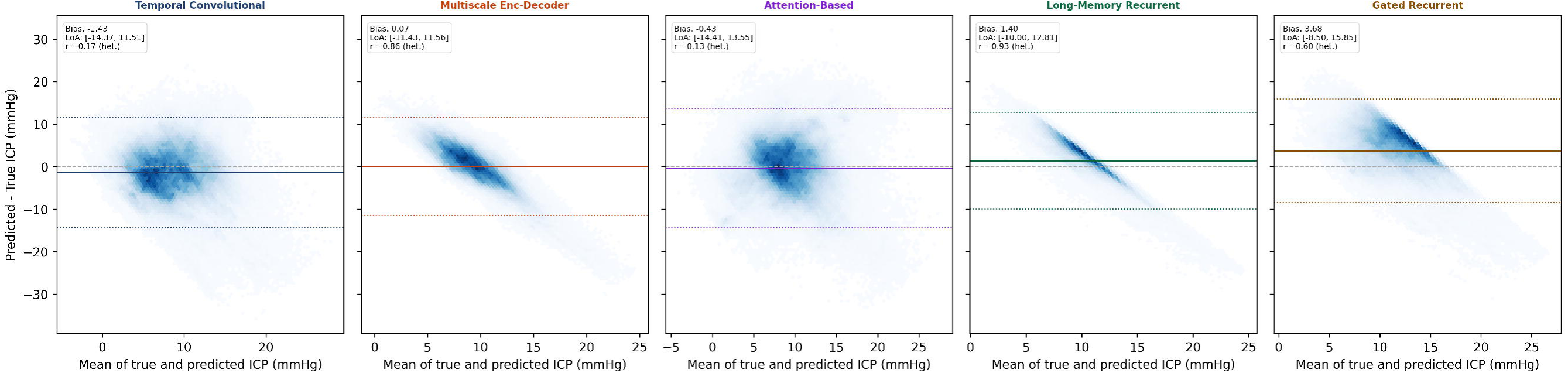
Agreement between predicted and true ICP across model configurations. Bland-Altman plots for the internally selected configuration of each model on the MIMIC-III matched validation cohort (n = 606,280 segments). Bias and 95% limits of agreement are shown. Heteroscedasticity was assessed via Pearson correlation between the mean and difference values (r, het.)

**Fig. 6.**
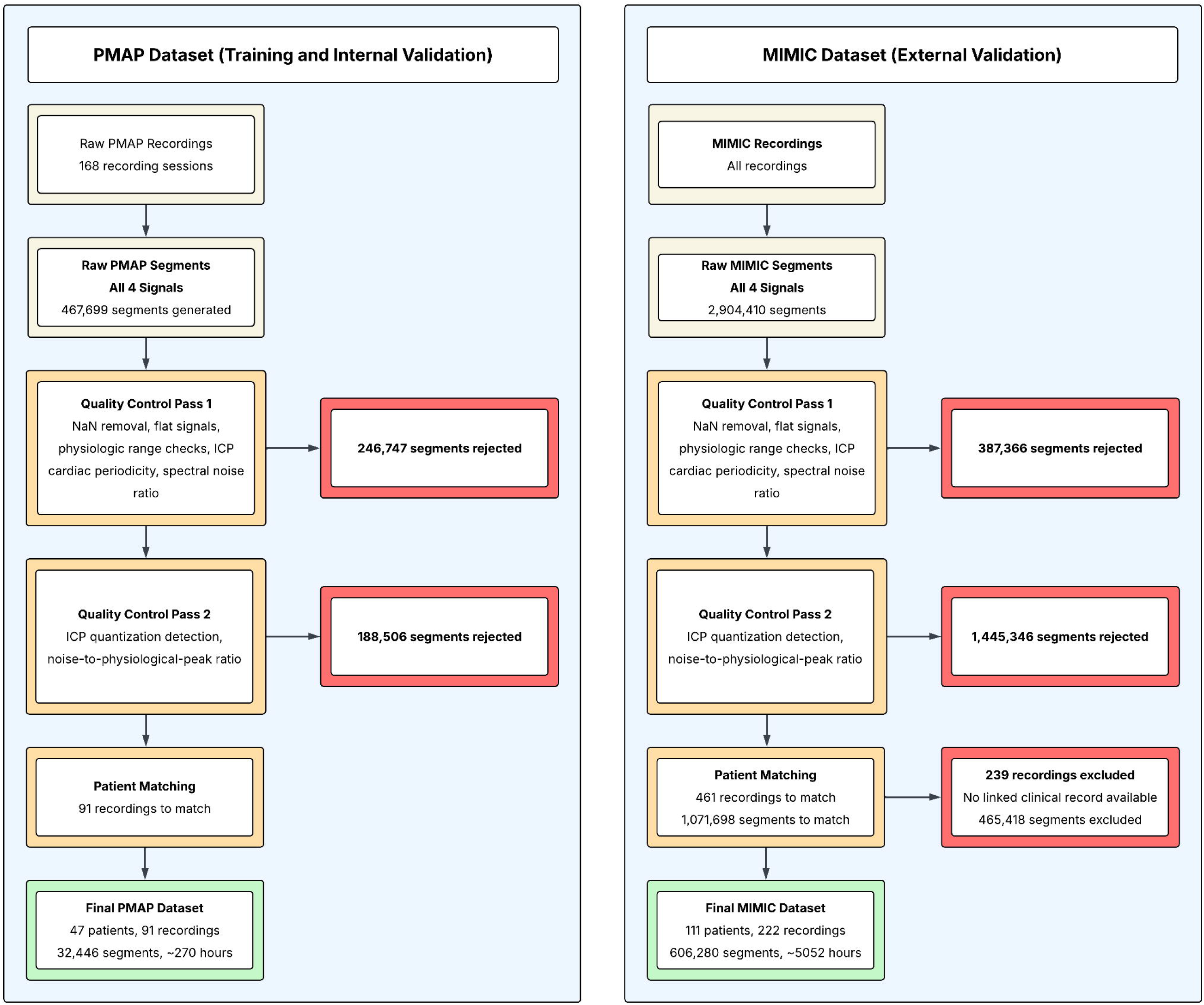
Patient inclusion and exclusion flowchart. Left panel: PMAP training cohort (Johns Hopkins Hospital). Right panel: MIMIC-III external validation cohort (Beth Israel Deaconess Medical Center). Quality control was applied in two passes: Pass 1 filtered segments on physiologic validity (NaN removal, range checks, flat signal detection, ICP cardiac periodicity, heart rate validity, spectral noise ratio); Pass 2 filtered on ICP morphology (quantization detection, noise-to-physiologic-peak ratio). For MIMIC-III, recordings surviving quality control were linked to clinical records via the PhysioNet matched subset.

### Declaration of Generative AI Usage

During the preparation and revision of this work, the authors used Anthropic’s Claude to improve the concision and clarity of the text. After using this service, the authors reviewed and edited the content as needed and take full responsibility for the content of the published article.

## Supporting information

All Supplementary Tables Here

## Data Availability

The MIMIC-III Waveform Database used for external validation is publicly available via PhysioNet. The PMAP training dataset is proprietary institutional data from Johns Hopkins Hospital and is not publicly available due to patient privacy constraints.

https://physionet.org/content/mimic3wdb/1.0/

## Statements and Declarations

### Data availability

The MIMIC-III Waveform Database used for external validation is publicly available via PhysioNet [22]. The PMAP training dataset is proprietary institutional data from Johns Hopkins Hospital and cannot be shared, including upon request, due to patient privacy constraints and institutional data governance restrictions. Queries regarding the dataset may be directed to the corresponding author.

### Code availability

The model architecture, hyperparameters, and training and inference code are available at *github.com/AnshGoyal1123/LCICM-ICP-Waveform-Generation* under a non-commercial academic research license; commercial use requires a license from Johns Hopkins Technology Ventures. The trained weights are available to the editors and reviewers during peer review and are available to academic investigators under a Model Use Agreement.

## Acknowledgements

The authors would like to thank Dr. Sofia Dias from the Department of Intensive Care at Unidade Local de Saúde São José, Lisbon, Portugal, for her contributions and insights into the clinical relevance of the project.

## Author Contributions

A.G. and V.Z. : conceptualization, methodology, software, formal analysis, data curation, visualization, writing – original draft, and writing – review & editing.

C.H. and R.D.S. : conceptualization, investigation, supervision, project administration, writing – original draft, and writing – review & editing.

All authors read and approved of the final manuscript.

## Ethics Approval

This study was approved by the Johns Hopkins Medicine Institutional Review Board (IRB00320184) and was performed in accordance with the ethical standards of the Declaration of Helsinki. The MIMIC-III Waveform Database is a publicly available, de-identified dataset; use of these data was exempt from additional IRB review.

## Consent to Participate

This study was conducted retrospectively using de-identified data. Informed consent was waived by the Johns Hopkins Medicine Institutional Review Board (IRB00320184) in view of the retrospective nature of the study. The MIMIC-III Waveform Database is fully de-identified; no additional consent was required.

