## Supplementary material for "Intracranial pressure waveform estimation using extracranial signals with deep learning": All Supplementary Tables Here

**Supplementary Information**

**Table of Contents**

**Supplementary Table 1.** Per-filter segment rejection counts (PMAP dataset)	2

**Supplementary Table 2.** Per-filter segment rejection counts (MIMIC-III dataset)	2

**Supplementary Table 3.** NaN source analysis: per-channel NaN burden among rejected segments.	3

**Supplementary Table 4.** NaN source analysis: contiguous gap length among rejected segments.	3

**Supplementary Table 5.** External performance: optimal external MAE and Pearson r per model	4

**Supplementary Table 6.** Pairwise Wilcoxon tests between models (external MAE)	5

**Supplementary Table 7.** Wilcoxon tests: early vs. late fusion MAE	6

**Supplementary Table 8.** Pairwise Wilcoxon tests between training objectives (early fusion)	7

**Supplementary Table 9.** Normalization ablation: normalized vs. unnormalized input performance	8

**Supplementary Table 10.** Sensitivity analysis: balanced-score correlation weighting (1.0×–3.0×)	9

**Supplementary Table 11.** Lag-adjusted cross-correlation and null-baseline correction	10

**Supplementary Table 12.** Dynamic time warping distance	11

**Supplementary Table 13.** External performance stratified by age, sex, and diagnosis (MAE)	12

**Supplementary Table 14.** External performance stratified by age, sex, and diagnosis (Pearson r)	13

**Supplementary Table 15.** Segment-weighted vs. patient-averaged MAE and Pearson r.	14

**Supplementary Table 16.** Architectural Characteristics of the Five Deep Learning Models	15

**Supplementary Table 17.** Beat-level peak morphology of reconstructed ICP waveforms	16

**Supplementary Figure 1.** Segment count distribution per patient (MIMIC-III cohort)	18

**Supplementary Figure 2.** Segment count distribution per patient (PMAP cohort)	19

**Supplementary Figure 3.** Predicted vs. reference P2/P1 amplitude ratio	20

| **Filter** | **Segments Rejected** | **% of Raw** |
| --- | --- | --- |
| **NaN (pre-resample)** | 177,958 | 38.1% |
| **ICP range** | 39,948 | 8.5% |
| **ABP range** | 17,244 | 3.7% |
| **Flat signal** | 11,539 | 2.5% |
| **Non-periodic ICP** | 57 | 0.01% |
| **HR invalid** | 1 | <0.01% |
| **Pass 1 total kept** | 220,952 | 47.2% |
| **Noise ratio** | 188,503 | 40.3% |
| **Unique values** | 3 | <0.01% |
| **Final kept** | 32,446 | 6.9% |

**Supplementary Table 1.** Per-filter segment rejection counts and percentages for the PMAP dataset.

| **Filter** | **Segments Rejected** | **% of Raw** |
| --- | --- | --- |
| **NaN (pre-resample)** | 228,090 | 7.9% |
| **ICP range** | 28,157 | 1.0% |
| **ABP range** | 40,514 | 1.4% |
| **Flat signal** | 87,290 | 3.0% |
| **Non-periodic ICP** | 2802 | 0.1% |
| **HR invalid** | 513 | 0.02% |
| **Pass 1 total kept** | 2,517,044 | 86.7% |
| **Noise ratio** | 942,719 | 32.5% |
| **Unique values** | 502,627 | 17.3% |
| **Pass 2 total kept** | 1,071,698 | 36.9% |
| **Patient info missing** | 465,418 | 16.0% |
| **Final kept** | 606,280 | 20.9% |

**Supplementary Table 2.** Per-filter segment rejection counts and percentages for the MIMIC-III Waveform Database. Note that 'Patient info missing' reflects segments excluded due to incomplete linkage between the waveform and clinical demographic databases.

| **Channel** | **PMAP (avg. % NaN)** | **MIMIC-III (avg. % NaN)** |
| --- | --- | --- |
| **ICP** | 12.2% | 13.4% |
| **ABP** | 91.7% | 2.6% |
| **PPG** | 0.8% | 0.5% |
| **ECG** | 0.003% | 1.1% |

**Supplementary Table 3.** NaN source analysis: per-channel NaN burden among rejected segments. Average percentage of NaN samples per channel among segments rejected for pre-resample NaN values in each cohort.

**Analysis:** The dominant NaN-contributing channel differs by cohort. In PMAP, ABP had the largest mean per-channel NaN fraction among rejected segments (91.7%), while in MIMIC-III, ABP is comparatively clean (2.6%) and ICP is instead the leading contributor (13.4%). This indicates that NaN-driven rejection reflects dataset- and protocol-specific factors rather than a fixed, universal signal-quality limitation.

| **Metric** | **PMAP** | **MIMIC-III** |
| --- | --- | --- |
| **Median max gap** | 30.0 s (full segment) | 0.21 s |
| **90th percentile max gap** | 30.0 s (full segment) | 8.2 s |
| **% of segments, max gap ≤10 samples** | 0.0% | 30.9% |
| **% of segments, max gap ≤50 samples** | 0.0% | 68.6% |

**Supplementary Table 4.** NaN source analysis: contiguous gap length among rejected segments. Maximum contiguous NaN run length per rejected segment, across all channels. n=177,958 rejected segments (PMAP), n=228,090 rejected segments (MIMIC-III).

**Analysis:** NaN gap structure differs sharply between cohorts. In MIMIC-III, gaps are short and right-skewed: up to 68.6% of rejected segments have a maximum gap of 50 samples (0.4 s) or fewer, suggesting short-window imputation could plausibly recover a substantial fraction of these segments. In PMAP, rejections are near-uniformly full-segment (median and 90th percentile both equal to the full 30-second window), consistent with structural recording gaps — for example, periods prior to arterial line placement — rather than brief transient dropout; small-gap imputation is therefore not applicable to the majority of PMAP's NaN-driven rejections. Imputation was not implemented in either cohort in the present study to maintain consistent preprocessing between datasets and avoid introducing interpolation-derived signal; short-gap interpolation for MIMIC-III specifically is a reasonable target for future preprocessing refinement.

| **Model** | **Lowest External MAE** | | | **Highest External r** | | |
| --- | --- | --- | --- | --- | --- | --- |
|  | **Config** | **MAE [95% CI]** | **r** | **Config** | **r [95% CI]** | **MAE** |
| **Multiscale Encoder-Decoder** | Late / WM | 4.266 [3.964, 4.616] | 0.586 | Early / BRR | 0.721 [0.679, 0.755] | 4.493 |
| **Long-Memory Recurrent** | Late / BRR | 4.330 [4.010, 4.698] | 0.493 | Late / WM | 0.656 [0.624, 0.686] | 4.353 |
| **Temporal Convolutional** | Late / WM | 4.358 [4.036, 4.736] | 0.599 | Early / WM | 0.663 [0.636, 0.687] | 4.847 |
| **Attention-Based** | Early / BRR | 4.946 [4.602, 5.390] | 0.722 | Early / BRR | 0.722 [0.683, 0.752] | 4.946 |
| **Gated Recurrent** | Late / WM | 4.276 [3.954, 4.633] | 0.619 | Late / WM | 0.619 [0.590, 0.644] | 4.276 |

**Supplementary Table 5. External Validation Performance (externally selected upper bound). Left:** configuration yielding optimal external MAE per model. **Right:** configuration yielding optimal external Pearson r. Because these configurations were chosen using the external data themselves, they represent an optimistic upper bound and are not the primary results; the primary (internally selected) results are in Table 2 of the main text. These results have been calculated on the MIMIC-III matched subset (n=606,280 segments, 111 patients).

**Analysis:** In the left panel ("Lowest External MAE"), late fusion was selected in four of the five models, suggesting that it more often supported external error minimization. In contrast, in the right panel ("Highest External r"), early fusion appeared in three of the five models, suggesting that it more often favored waveform-shape agreement. The waveform-morphology objective (WM) appeared most often in the lowest-MAE and was the objective used in all five internally selected configurations reported in Table 2 of the main text. Meanwhile, BRR appeared more often in the highest-correlation settings. WRR did not appear as the lowest-MAE configuration for any model.

| **Model A** | **Config A** | **MAE A** | **Model B** | **Config B** | **MAE B** | **p (corrected)** | **Significant** |
| --- | --- | --- | --- | --- | --- | --- | --- |
| **MED** | Late/WM | 4.266 | **LMR** | Early/WM | 4.637 | 0.250 | **No** |
| **MED** | Late/WM | 4.266 | **TC** | Early/WM | 4.847 | 0.007 | **Yes** |
| **MED** | Late/WM | 4.266 | **AB** | Early/WM | 5.285 | <0.001 | **Yes** |
| **MED** | Late/WM | 4.266 | **GR** | Early/WM | 5.899 | <0.001 | **Yes** |
| **LMR** | Early/WM | 4.637 | **TC** | Early/WM | 4.847 | 0.681 | **No** |
| **LMR** | Early/WM | 4.637 | **AB** | Early/WM | 5.285 | 0.008 | **Yes** |
| **LMR** | Early/WM | 4.637 | **GR** | Early/WM | 5.899 | <0.001 | **Yes** |
| **TC** | Early/WM | 4.847 | **AB** | Early/WM | 5.285 | 0.217 | **No** |
| **TC** | Early/WM | 4.847 | **GR** | Early/WM | 5.899 | 0.002 | **Yes** |
| **AB** | Early/WM | 5.285 | **GR** | Early/WM | 5.899 | 0.392 | **No** |

**Supplementary Table 6.** Pairwise Wilcoxon signed-rank tests between models on external validation MAE, using the internally selected configuration per model (Table 2 of the main text), paired on per-patient mean MAE. Bonferroni correction applied (k=10). TC = Temporal Convolutional; MED = Multiscale Encoder-Decoder; AB = Attention-Based; LMR = Long-Memory Recurrent; GR = Gated Recurrent. WM = waveform-morphology objective; BRR = baseline robust regression; WRR = weighted robust regression. n=111 patients (606,280 segments).

| **Model** | **Objective** | **MAE Early** | **MAE Late** | **p (raw)** | **Significant** |
| --- | --- | --- | --- | --- | --- |
| **MED** | BRR | 4.493 | 4.595 | 0.282 | No |
| **MED** | WRR | 4.630 | 4.587 | 0.116 | No |
| **MED** | WM | 4.490 | 4.266 | 0.067 | No |
| **LMR** | BRR | 4.872 | 4.330 | 0.002 | Yes |
| **LMR** | WRR | 4.535 | 4.389 | 0.043 | Yes |
| **LMR** | WM | 4.637 | 4.353 | 0.006 | Yes |
| **TC** | BRR | 5.127 | 4.490 | 0.004 | Yes |
| **TC** | WRR | 5.090 | 4.602 | 0.013 | Yes |
| **TC** | WM | 4.847 | 4.358 | 0.038 | Yes |
| **AB** | BRR | 4.946 | 5.091 | 0.972 | No |
| **AB** | WRR | 5.159 | 5.044 | 0.548 | No |
| **AB** | WM | 5.285 | 5.186 | 0.888 | No |
| **GR** | BRR | 5.238 | 4.439 | <0.001 | Yes |
| **GR** | WRR | 4.782 | 5.605 | <0.001 | Yes |
| **GR** | WM | 5.899 | 4.276 | <0.001 | Yes |

**Supplementary Table 7.** Wilcoxon signed-rank tests comparing early vs. late fusion MAE for each model-objective combination. Tests were paired on per-patient mean MAE (n=111 patients); MAE columns are segment-weighted means over 606,280 segments, as in Table 2. No correction applied. Abbreviations as in Supplementary Table 6.

| **Model** | **Objective A** | **MAE A** | **Objective B** | **MAE B** | **p (corrected)** | **Significant** |
| --- | --- | --- | --- | --- | --- | --- |
| **MED** | BRR | 4.493 | WRR | 4.630 | 0.060 | No |
| **MED** | BRR | 4.493 | WM | 4.490 | 0.410 | No |
| **MED** | WRR | 4.630 | WM | 4.490 | 0.123 | No |
| **LMR** | BRR | 4.872 | WRR | 4.535 | 0.690 | No |
| **LMR** | BRR | 4.872 | WM | 4.637 | <0.001 | Yes |
| **LMR** | WRR | 4.535 | WM | 4.637 | 1.000 | No |
| **TC** | BRR | 5.127 | WRR | 5.090 | 1.000 | No |
| **TC** | BRR | 5.127 | WM | 4.847 | 0.224 | No |
| **TC** | WRR | 5.090 | WM | 4.847 | 0.122 | No |
| **AB** | BRR | 4.946 | WRR | 5.159 | 0.905 | No |
| **AB** | BRR | 4.946 | WM | 5.285 | 0.810 | No |
| **AB** | WRR | 5.159 | WM | 5.285 | 1.000 | No |
| **GR** | BRR | 5.238 | WRR | 4.782 | 0.002 | Yes |
| **GR** | BRR | 5.238 | WM | 5.899 | 0.653 | No |
| **GR** | WRR | 4.782 | WM | 5.899 | <0.001 | Yes |

**Supplementary Table 8.** Pairwise Wilcoxon signed-rank tests between training objectives within each model (early fusion). Tests were paired on per-patient mean MAE (n=111 patients); MAE columns are segment-weighted means over 606,280 segments, as in Table 2. Bonferroni correction applied (k=3 per model); the p-value shown is the corrected value. Abbreviations as in Supplementary Table 6.

| **Model** | **Fusion** | **Regular MAE** | **Norm MAE [95% CI]** | **ΔMAE** | **Regular r** | **Norm r [95% CI]** | **Δr** |
| --- | --- | --- | --- | --- | --- | --- | --- |
| **MED** | Early | 4.493 | 6.388 [5.789, 7.069] | +1.895 | 0.721 | 0.320 [0.288, 0.352] | −0.401 |
| **LMR** | Late | 4.353 | 4.449 [4.074, 4.869] | +0.096 | 0.656 | 0.639 [0.605, 0.672] | −0.017 |
| **TC** | Late | 4.358 | 4.439 [4.064, 4.867] | +0.081 | 0.599 | 0.604 [0.569, 0.636] | +0.005 |
| **AB** | Early | 4.946 | 5.510 [5.133, 5.973] | +0.564 | 0.722 | 0.641 [0.610, 0.670] | −0.081 |
| **GR** | Late | 4.276 | 4.424 [4.041, 4.858] | +0.148 | 0.619 | 0.645 [0.610, 0.679] | +0.026 |

**Supplementary Table 9.** Normalization ablation tested on the externally selected configurations presented in Table 3 of the main text. Normalization was performed per-segment and consisted of a per-channel z-scoring of ABP, PPG, and ECG. Normalization was applied at training and at inference. n=606,280 segments, 111 patients.

**Analysis:** Late fusion models retrained with per-segment, per-channel z-score normalization applied to input signals (ABP/PPG/ECG) were minimally affected relative to the unnormalized values reported in Table 3 of the main text. Early fusion models, however, were substantially degraded, most severely the multiscale encoder-decoder. This is an expected result, considering that normalization of signals flattens all relative amplitude information between channels, which the early fusion models likely used to make their predictions. Since late fusion models do not see relative amplitude differences across channels, they were less severely affected by normalization. Regardless, normalization did not improve MAE in any configuration tested and changed r only marginally for the late-fusion models (−0.017 to +0.026), which supports the choice to avoid it. This ablation was run on the externally selected configurations (Table 3 of the main text) and was not repeated for the internally selected configurations reported in Table 2.

| **Architecture** | **w = 1.0** | **w = 1.5 (reported)** | **w = 2.0** | **w = 3.0** |
| --- | --- | --- | --- | --- |
| **MED** | **Late/BRR**  MAE: 4.595  r: 0.432 | **Late/WM**  MAE: 4.266  r: 0.586 | **Late/WM**  MAE: 4.266  r: 0.586 | **Late/WM**  MAE: 4.266  r: 0.586 |
| **LMR** | **Early/WM**  MAE: 4.637  r: 0.585 | **Early/WM**  MAE: 4.637  r: 0.585 | **Early/WM**  MAE: 4.637  r: 0.585 | **Early/WM**  MAE: 4.637  r: 0.585 |
| **TC** | **Early/WM**  MAE: 4.847  r: 0.663 | **Early/WM**  MAE: 4.847  r: 0.663 | **Early/WM**  MAE: 4.847  r: 0.663 | **Early/WM**  MAE: 4.847  r: 0.663 |
| **AB** | **Early/WM**  MAE: 5.285  r: 0.719 | **Early/WM**  MAE: 5.285  r: 0.719 | **Early/WM**  MAE: 5.285  r: 0.719 | **Early/WM**  MAE: 5.285  r: 0.719 |
| **GR** | **Early/WM**  MAE: 5.899  r: 0.601 | **Early/WM**  MAE: 5.899  r: 0.601 | **Early/WM**  MAE: 5.899  r: 0.601 | **Early/WM**  MAE: 5.899  r: 0.601 |

**Supplementary Table 10.** Internally selected configuration per architecture under balanced-score correlation weights ranging from 1.0× to 3.0× (selection performed on PMAP validation data; the primary analysis uses 1.5×). Reported MAE and r are the external (MIMIC-III) performance of each selected configuration. n=606,280 segments, 111 patients.

**Analysis:** The internally selected configuration was identical across the full tested range for four of five architectures (TC, AB, LMR, GR), indicating that the reported weighting choice had no effect on which configuration was ultimately reported for these models. MED was the only sensitive architecture: at w=1.0, the unweighted balanced score selected the baseline robust regression objective instead of the waveform-morphology objective (both late fusion). That alternative was worse on both external metrics (MAE 4.595 vs 4.266 mmHg; r=0.432 vs 0.586), so the switch would not have improved external performance. Because the weighting only changed one internal selection, and only at the unweighted setting, the choice of w=1.5 is not driving the reported results. For reference, the externally selected (upper-bound) configurations were more sensitive: they changed for two architectures (TC at w=3.0; MED at w=1.0), consistent with selection on a single external dataset being less stable than selection on the internal validation data.

| **Model** | **Zero-lag r [95% CI]** | **Lag-adj r [95% CI]** | **Mean optimal lag (ms)** | **Corrected improvement** |
| --- | --- | --- | --- | --- |
| **MED** | 0.586 [0.551, 0.618] | 0.695 [0.667, 0.723] | +1.2 | +0.015 |
| **LMR** | 0.585 [0.529, 0.631] | 0.773 [0.745, 0.799] | +62.1 | +0.098 |
| **TC** | 0.663 [0.636, 0.687] | 0.713 [0.692, 0.732] | +14.4 | −0.034 |
| **AB** | 0.719 [0.682, 0.749] | 0.793 [0.772, 0.812] | +12.4 | −0.014 |
| **GR** | 0.601 [0.544, 0.647] | 0.759 [0.727, 0.788] | +39.7 | +0.072 |

**Supplementary Table 11.** Lag-adjusted cross-correlation and null-baseline correction. Lag search window ±75 samples (±0.6 s at 125 Hz). Corrected improvement is the observed lag-adjustment gain minus the mean improvement obtained from 5,000 shuffled true/predicted pairs subjected to the same lag search, controlling for the possibility that searching many candidate lags inflates correlation by chance alone. The shuffled null pairs were drawn from the saved patient-stratified waveform subset of approximately 10,000 segments rather than from all segments. n=606,280 segments, 111 patients.

**Analysis:** Raw lag-adjustment improvement was positive for all five architectures (+0.05 to +0.19). After correction against the null baseline, the corrected improvement was at or below zero for TC (−0.034) and AB (−0.014) and negligibly positive for MED (+0.015), indicating that for these three architectures the apparent gain is no greater than lag search alone produces on shuffled, unrelated signal pairs. LMR and GR differed: their corrected improvements remained positive (+0.098 and +0.072) and they had the largest mean optimal lags (62 and 40 ms, versus 14.4 ms or less for the other three). For these two models, part of the gain from lag adjustment (zero-lag r of 0.585 and 0.601 rising to 0.773 and 0.759) reflects a genuine timing offset in the predicted waveform and not just a lag-search selection effect, and their reported Pearson r values are therefore modestly deflated by small temporal misalignment. For TC, MED, and AB, the reported r values are not meaningfully deflated by timing misalignment.

| **Model** | **Shape-normalized DTW** | **95% CI** | **Amplitude-preserving DTW (mmHg)** | **95% CI** |
| --- | --- | --- | --- | --- |
| **AB** | 20.729 | [19.602, 21.934] | 4.876 | [4.471, 5.341] |
| **TC** | 23.856 | [22.904, 24.795] | 4.168 | [3.748, 4.696] |
| **GR** | 24.275 | [22.193, 26.454] | 5.107 | [4.644, 5.642] |
| **LMR** | 24.583 | [22.817, 26.489] | 4.354 | [3.983, 4.761] |
| **MED** | 25.083 | [23.659, 26.550] | 4.231 | [3.912, 4.611] |

**Supplementary Table 12.** Dynamic time warping distance calculated for the 5 internally selected configurations from Table 2. DTW distance computed with a bounded warping window (±75 samples, ±0.6 s at 125 Hz) under two configurations. Shape-normalized DTW is computed on z-score normalized signals, isolating waveform-shape agreement independent of absolute amplitude (unitless; lower values indicate closer shape agreement). Amplitude-preserving DTW is computed on raw (mmHg) values, normalized by √T for per-sample interpretability, retaining absolute error magnitude. n=606,280 segments, 111 patients.

**Analysis:** Shape-normalized DTW was lowest (tightest waveform agreement) for AB (20.7), followed by TC (23.9), GR (24.3), LMR (24.6), and MED (25.1); the confidence intervals for the latter four overlap substantially. This ordering tracks Pearson r rather than MAE: AB has the highest external r (0.719), whereas MED has the lowest MAE but the largest shape-normalized DTW. Amplitude-preserving DTW (per-sample, mmHg) was lowest for TC (4.17) and MED (4.23), followed by LMR (4.35), AB (4.88), and GR (5.11), and so largely follows the MAE ranking, as expected since both retain absolute error magnitude. Together with Supplementary Table 11, these results show that neither timing misalignment nor amplitude error alone explains architecture differences, and that MAE and Pearson r capture most of the information that DTW assesses in our case.

| **Patient Group (n)** | **MED** | **LMR** | **TC** | **AB** | **GR** |
| --- | --- | --- | --- | --- | --- |
| **SAH/Aneurysm (35)** | **3.88**  [3.61, 4.17] | **4.36**  [3.86, 4.91] | **4.24**  [3.94, 4.58] | **4.62**  [4.25, 5.03] | **5.95**  [5.25, 6.68] |
| **Brain tumor/mass (9)** | **4.55**  [3.17, 5.54] | **4.97**  [3.58, 5.87] | **3.94**  [3.36, 4.25] | **4.02**  [3.32, 4.37] | **6.24**  [4.07, 7.53] |
| **Other ICH (38)** | **4.42**  [3.78, 5.27] | **4.82**  [4.09, 5.69] | **5.11**  [4.40, 6.21] | **5.62**  [4.90, 6.58] | **5.85**  [5.01, 6.92] |
| **Trauma (5)** | **5.29**  [4.31, 10.18] | **4.58**  [4.25, 8.80] | **6.93**  [4.37, 9.76] | **7.41**  [5.62, 13.27] | **3.91**  [3.71, 7.27] |
| **Other neuro (9)** | **3.82**  [3.10, 4.73] | **4.08**  [3.71, 5.49] | **4.57**  [4.06, 7.10] | **5.55**  [3.97, 9.09] | **5.51**  [4.74, 7.24] |
| **Other medical (15)** | **5.30**  [4.66, 5.92] | **5.30**  [4.75, 6.09] | **6.68**  [4.92, 8.63] | **6.80**  [5.68, 8.24] | **6.10**  [5.07, 8.01] |
| **Male (46)** | **4.42**  [3.87, 5.29] | **4.58**  [3.96, 5.46] | **5.01**  [4.22, 6.17] | **5.59**  [4.93, 6.47] | **5.78**  [4.99, 6.77] |
| **Female (65)** | **4.17**  [3.85, 4.48] | **4.67**  [4.28, 5.07] | **4.75**  [4.37, 5.25] | **5.09**  [4.63, 5.60] | **5.98**  [5.44, 6.56] |
| **Age Group: 18-39 (12)** | **4.50**  [4.06, 5.64] | **3.88**  [3.56, 4.83] | **4.27**  [3.78, 5.75] | **5.27**  [4.41, 7.74] | **4.01**  [3.69, 4.87] |
| **Age Group: 40-59 (41)** | **4.32**  [3.88, 4.83] | **4.38**  [3.95, 4.87] | **4.90**  [4.26, 5.99] | **5.54**  [4.84, 6.23] | **5.66**  [5.07, 6.42] |
| **Age Group: 60-79 (47)** | **4.34**  [3.89, 4.90] | **4.93**  [4.39, 5.52] | **4.99**  [4.41, 5.78] | **5.17**  [4.69, 5.78] | **6.33**  [5.72, 7.00] |
| **Age Group: ≥80 (11)** | **3.56**  [3.34, 4.16] | **4.28**  [3.73, 5.18] | **4.45**  [3.70, 5.06] | **5.31**  [3.76, 6.72] | **5.69**  [4.39, 7.50] |
| **All patients (111)** | **4.27**  [3.96, 4.62] | **4.64**  [4.28, 5.03] | **4.85**  [4.44, 5.35] | **5.29**  [4.91, 5.72] | **5.90**  [5.43, 6.43] |

**Supplementary Table 13.** External performance stratified by age, sex, and primary diagnosis (MAE, mmHg [95% CI]). 95% CIs from 10,000 patient-cluster bootstrap resamples using each architecture's internally selected configuration from Table 2. n=606,280 segments across 111 patients.

**Analysis:** MAE varied by primary diagnosis, and no single architecture performed best across all categories. The two largest subgroups, SAH/aneurysm (n=35) and other ICH (n=38), together comprising 66% of the cohort, produced MAE point estimates within 0.7 mmHg of the overall cohort value for all five architectures, with comparatively narrow confidence intervals, indicating that Table 2's headline results are not disproportionately driven by any one diagnostic subpopulation. The trauma subgroup (n=5) had the widest MAE confidence interval for all five architectures and the highest MAE point estimate for two of them (attention-based and temporal convolutional), but the lowest for the gated recurrent model, and should be treated as exploratory given the small sample. These results, considered alongside Supplementary Table 14, indicate that diagnosis-related performance variation exists but does not overturn the overall pattern of results reported in Table 2. Age-stratified MAE showed no consistent pattern: the lowest point estimate was in the 18–39 group for three architectures (TC, LMR, GR), the ≥80 group for MED, and the 60–79 group for AB, and confidence intervals overlapped substantially across age groups. Performance was broadly similar between male and female patients, with overlapping confidence intervals across all architectures; female patients had lower MAE point estimates for three of five models (TC, MED, AB) and higher for two (LMR, GR).

| **Patient Group (n)** | **MED** | **LMR** | **TC** | **AB** | **GR** |
| --- | --- | --- | --- | --- | --- |
| **SAH/Aneurysm (35)** | **0.61**  [0.57, 0.66] | **0.63**  [0.56, 0.68] | **0.68**  [0.66, 0.70] | **0.74**  [0.70, 0.77] | **0.63**  [0.57, 0.69] |
| **Brain tumor/mass (9)** | **0.54**  [0.41, 0.59] | **0.65**  [0.50, 0.70] | **0.65**  [0.52, 0.71] | **0.74**  [0.62, 0.78] | **0.67**  [0.52, 0.73] |
| **Other ICH (38)** | **0.56**  [0.49, 0.63] | **0.57**  [0.50, 0.65] | **0.64**  [0.60, 0.66] | **0.71**  [0.67, 0.74] | **0.60**  [0.53, 0.67] |
| **Trauma (5)** | **0.49**  [0.44, 0.49] | **0.27**  [0.26, 0.50] | **0.52**  [0.51, 0.65] | **0.54**  [0.50, 0.59] | **0.37**  [0.36, 0.64] |
| **Other neuro (9)** | **0.59**  [0.33, 0.64] | **0.64**  [0.59, 0.69] | **0.77**  [0.66, 0.79] | **0.80**  [0.73, 0.82] | **0.65**  [0.58, 0.72] |
| **Other medical (15)** | **0.60**  [0.51, 0.66] | **0.42**  [0.20, 0.64] | **0.61**  [0.50, 0.73] | **0.62**  [0.44, 0.80] | **0.43**  [0.19, 0.67] |
| **Male (46)** | **0.55**  [0.48, 0.61] | **0.52**  [0.47, 0.56] | **0.66**  [0.61, 0.70] | **0.71**  [0.67, 0.75] | **0.55**  [0.50, 0.59] |
| **Female (65)** | **0.61**  [0.58, 0.64] | **0.62**  [0.54, 0.69] | **0.67**  [0.63, 0.69] | **0.72**  [0.67, 0.76] | **0.63**  [0.55, 0.70] |
| **Age Group: 18-39 (12)** | **0.46**  [0.41, 0.60] | **0.46**  [0.33, 0.54] | **0.59**  [0.48, 0.66] | **0.62**  [0.55, 0.69] | **0.46**  [0.35, 0.54] |
| **Age Group: 40-59 (41)** | **0.59**  [0.53, 0.62] | **0.52**  [0.37, 0.62] | **0.66**  [0.57, 0.72] | **0.67**  [0.55, 0.76] | **0.52**  [0.37, 0.63] |
| **Age Group: 60-79 (47)** | **0.59**  [0.55, 0.64] | **0.63**  [0.57, 0.69] | **0.67**  [0.65, 0.70] | **0.75**  [0.72, 0.78] | **0.66**  [0.60, 0.70] |
| **Age Group: ≥80 (11)** | **0.65**  [0.56, 0.70] | **0.58**  [0.53, 0.62] | **0.69**  [0.65, 0.74] | **0.75**  [0.71, 0.77] | **0.60**  [0.53, 0.65] |
| **All patients (111)** | **0.59**  [0.55, 0.62] | **0.58**  [0.53, 0.63] | **0.66**  [0.64, 0.69] | **0.72**  [0.68, 0.75] | **0.60**  [0.54, 0.65] |

**Supplementary Table 14.** External performance stratified by age, sex, and primary diagnosis (Pearson r [95% CI]). 95% CIs from 10,000 patient-cluster bootstrap resamples, computed identically to Supplementary Table 13. n=606,280 segments across 111 patients.

**Analysis:** Similarly to the MAE results, correlation for the two largest subgroups most closely tracked the headline numbers presented in Table 2 (SAH/aneurysm r=0.61–0.74 and other ICH r=0.56–0.71, versus overall r=0.58–0.72). The trauma subgroup showed the lowest correlation for all five architectures (r=0.27–0.54), but this result is influenced by the smallest sample size of the analysis (n=5) and should not be used as anything but exploratory. The highest-correlation subgroup varied by architecture: the “other neuro” subgroup (9 patients) was highest for the attention-based (r=0.80) and temporal convolutional (r=0.77) models, whereas the brain tumor subgroup (9 patients) was highest for the two recurrent models and SAH/aneurysm for the multiscale encoder-decoder, and each of these is influenced by small sample sizes. Taken together, Supplementary Tables 13–14 show that no architecture is uniformly optimal across diagnostic categories and that estimates for smaller subgroups, particularly trauma, should be interpreted with appropriate caution given wide confidence intervals. Age-stratified correlation was lowest in the 18–39 group for all five architectures and highest in the 60–79 or ≥80 group (e.g., TC r=0.59 in the 18–39 group versus 0.69 in the ≥80 group), although confidence intervals generally overlapped and the youngest and oldest groups are small (12 and 11 patients). Female patients had higher correlation point estimates than male patients across all five architectures, although confidence intervals generally overlapped between sex groups.

| **Model** | **Config** | **Segment**  **Weighted**  **MAE**  **(mean ± SD)** | **Patient**  **Averaged**  **MAE**  **(mean ± SD)** | **Patient**  **Averaged**  **MAE**  **(median [IQR])** | **Segment**  **Weighted**  **r**  **(mean ± SD)** | **Patient**  **Averaged**  **r**  **(mean ± SD)** | **Patient**  **Averaged**  **r**  **(median [IQR])** |
| --- | --- | --- | --- | --- | --- | --- | --- |
| **MED** | Late/WM | **4.266**  ± 2.815 | **5.284**  ± 3.401 | **4.077**  [3.409, 5.380] | **0.586**  ± 0.205 | **0.498**  ± 0.208 | **0.546**  [0.417, 0.647] |
| **LMR** | Early/WM | **4.637**  ± 2.892 | **5.506**  ± 3.349 | **4.501**  [3.379, 6.018] | **0.585**  ± 0.245 | **0.474**  ± 0.228 | **0.518**  [0.367, 0.639] |
| **TC** | Early/WM | **4.847**  ± 3.514 | **5.831**  ± 3.546 | **4.652**  [3.946, 6.142] | **0.663**  ± 0.181 | **0.566**  ± 0.209 | **0.635**  [0.478, 0.698] |
| **AB** | Early/WM | **5.285**  ± 3.721 | **6.174**  ± 3.437 | **5.014**  [4.135, 6.826] | **0.719**  ± 0.211 | **0.606**  ± 0.232 | **0.692**  [0.531, 0.769] |
| **GR** | Early/WM | **5.899**  ± 3.425 | **6.730**  ± 3.461 | **5.866**  [4.614, 7.808] | **0.601**  ± 0.242 | **0.490**  ± 0.243 | **0.524**  [0.411, 0.648] |

**Supplementary Table 15.** Segment-weighted vs. patient-averaged MAE and Pearson r. Segment-weighted values are computed by averaging per-segment MAE/r across all 606,280 external validation segments (identical methodology to Table 2). Patient-averaged values are computed by first averaging per-segment MAE/r within each of the 111 patients, then summarizing across patients. n=606,280 segments, 111 patients.

**Analysis:** Patient-averaged mean MAE (5.28–6.73 mmHg) exceeded the segment-weighted value for all five architectures and exceeded the 5-mmHg clinically motivated MAE benchmark for all five, while the patient-averaged median (4.08–5.87 mmHg) remained below the benchmark for three of five architectures (TC, MED, LMR) and closely tracked the segment-weighted mean, indicating a right-skewed distribution of per-patient MAE driven by a subset of patients with disproportionately high error. This divergence was only weakly and inconsistently associated with a patient's representation of hypertensive-range segments (see Results), and the ICP-range-dependent underprediction reported in the stratified Bland-Altman analysis (Table 4, Fig. 5) is a more consistent characterization of the limitation than any single patient subset.

| **Model Architecture** | **Architecture Description** |
| --- | --- |
| **Temporal Convolutional** | Initial 1D convolution with 64 filters, followed by six residual dilated-convolution blocks with dilation rates 1, 2, 4, 8, 16, and 32. Each block contains two 1D convolutions with 64 filters and kernel size 5. A final 64-filter convolution and 1-channel output convolution produce the ICP waveform. |
| **Multiscale Encoder-Decoder** | Four-level 1D encoder-decoder with skip connections. Encoder filter widths increase from 16→32→64→128, with two kernel-size-3 convolutions per level and ×2 max pooling. The bottleneck uses 256 filters. The decoder symmetrically upsamples through 128→64→32→16 filters with concatenated encoder skip connections, followed by a 1-channel linear output layer. |
| **Attention-Based** | Initial strided 1D convolution with 128 filters followed by two Transformer blocks. Each block uses 128-dimensional representations, 4 attention heads, and a 256-unit feed-forward layer with residual connections and layer normalization. The representation is reduced to 64 dimensions, upsampled to the original temporal resolution, and mapped to a 1-channel ICP output. |
| **Long-Memory Recurrent** | Three stacked LSTM layers with 256, 128, and 64 hidden units, respectively, each returning the full temporal sequence, followed by a 1-unit dense output layer. |
| **Gated Recurrent** | Three stacked GRU layers with 256, 128, and 64 hidden units, respectively, each returning the full temporal sequence, followed by a 1-unit dense output layer. |

**Supplementary Table 16.** This table summarizes the principal architectural characteristics of the five sequence-to-sequence models evaluated in this study. For each model, the table describes the major computational components, including convolutional or recurrent layer structure, filter or hidden-unit dimensions, dilation or attention configuration where applicable, skip connections, and the final waveform output layer.

| **Metric** | **MED** | **LMR** | **TC** | **AB** | **GR** |
| --- | --- | --- | --- | --- | --- |
| P1–P2 pair detected | 0.030 [0.022, 0.042] | 0.012 [0.008, 0.016] | 0.749 [0.700, 0.790] | 0.416 [0.336, 0.491] | 0.171 [0.095, 0.245] |
| P1-dominance accuracy | 0.679 [0.618, 0.726] | 0.727 [0.650, 0.780] | 0.673 [0.625, 0.711] | 0.751 [0.685, 0.804] | 0.874 [0.801, 0.923] |
| *Majority-class baseline* | 0.774 [0.710, 0.821] | 0.702 [0.575, 0.791] | 0.799 [0.750, 0.838] | 0.824 [0.770, 0.865] | 0.863 [0.776, 0.920] |
| P1-dominance kappa | 0.170 [0.094, 0.243] | 0.361 [0.255, 0.447] | 0.106 [0.061, 0.145] | 0.147 [0.054, 0.230] | 0.316 [0.174, 0.501] |
| P2/P1 ratio r | 0.145 [0.087, 0.212] | 0.289 [0.215, 0.362] | 0.143 [0.098, 0.191] | 0.279 [0.219, 0.335] | 0.402 [0.289, 0.518] |
| *Shuffled-pair null r* | 0.028 [0.004, 0.052] | 0.124 [0.072, 0.177] | 0.030 [0.023, 0.037] | 0.038 [0.029, 0.048] | 0.070 [0.057, 0.083] |
| P2/P1 ratio MAE | 0.435 [0.377, 0.493] | 0.457 [0.397, 0.543] | 0.459 [0.391, 0.556] | 0.344 [0.300, 0.405] | 0.356 [0.310, 0.416] |
| *Constant baseline* | 0.352 [0.293, 0.418] | 0.382 [0.301, 0.501] | 0.299 [0.251, 0.362] | 0.279 [0.230, 0.352] | 0.243 [0.202, 0.326] |
| P1 latency MAE (ms) | 65.0 [53.3, 79.0] | 67.9 [60.1, 80.5] | 59.2 [52.5, 67.3] | 56.6 [50.3, 64.7] | 71.1 [64.4, 81.4] |
| *Constant baseline (ms)* | 57.9 [42.5, 74.1] | 45.1 [35.5, 60.4] | 38.5 [29.0, 50.9] | 36.2 [27.1, 49.1] | 24.6 [20.6, 32.9] |
| P1–P2 interval MAE (ms) | 149.7 [120.7, 181.8] | 151.5 [128.3, 178.7] | 87.9 [82.4, 95.3] | 89.6 [79.9, 101.3] | 79.5 [66.8, 97.8] |
| *Constant baseline (ms)* | 118.3 [89.8, 148.4] | 178.3 [129.7, 244.1] | 69.2 [60.4, 81.8] | 72.6 [60.6, 90.2] | 73.7 [60.6, 95.3] |
| P3 sensitivity | 0.002 [0.001, 0.003] | 0.001 [0.000, 0.001] | 0.492 [0.426, 0.547] | 0.081 [0.059, 0.104] | 0.011 [0.006, 0.018] |
| P3 specificity | 0.999 [0.998, 1.000] | 1.000 [1.000, 1.000] | 0.572 [0.534, 0.607] | 0.958 [0.945, 0.969] | 0.999 [0.998, 0.999] |

**Supplementary Table 17.** Beat-level peak morphology of reconstructed ICP waveforms (MIMIC-III external validation; internally selected configurations). Analysis used a subset of 10,030 external segments from all 111 patients. Beats were delimited at troughs of the reference ICP signal (0.4–1.5 s, amplitude ≥0.5 mmHg), the same boundaries were applied to the prediction, and P1, P2 and P3 were taken as the first three local maxima of each beat (prominence ≥10% of the reference beat amplitude, minimum spacing 64 ms). P1 and P2 were identifiable in 42.6% of reference beats (95% CI 33.6–52.4%) and P3 in 13.3% (9.2–18.2%). Peak metrics are computed on reference beats with identifiable peaks; "P1–P2 pair detected" is the proportion of reference beats with identifiable P1 and P2 in which the prediction also contained both peaks. Shaded rows give the corresponding baselines ("Constant baseline" is the constant-value baseline described below). The constant-value baseline predicts the median reference value of each metric for every beat; the shuffled-pair null is the P2/P1 ratio correlation after randomly shuffling predicted ratios across beats within each patient; P1-dominance is P1 > P2, and its majority-class baseline is the accuracy of always predicting the more common class. Values are mean [95% CI] from 10,000 patient-level bootstrap resamples. MED, multiscale encoder-decoder; LMR, long-memory recurrent; TC, temporal convolutional; AB, attention-based; GR, gated recurrent.

**Analysis:** The multiscale encoder-decoder and long-memory recurrent models produced a distinguishable P1–P2 pair in only 3.0% and 1.2% of reference beats that had one, compared with 74.9% (TC), 41.6% (AB) and 17.1% (GR). Where peaks were detected, P2/P1 ratio correlation (0.14–0.40) exceeded the shuffled-pair null (0.03–0.12), but P2/P1 ratio and P1 latency errors were higher than the constant-value baseline for every model, and P1–P2 interval error was higher for four of five (the exception, LMR, rests on very few detected pairs). P1-dominance accuracy was below the majority-class baseline for TC, MED and AB and only marginally above it for LMR and GR (kappa 0.36 and 0.32), and P3 sensitivity was at most 0.08 for all models except TC (0.49 sensitivity with 0.57 specificity, near chance). Taken together, the models recover mean pressure level and the dominant pulsatile component but not beat-level peak configuration. Because inter-signal timing was not corrected, latency errors may be partly attributable to timing offsets, although amplitude-ratio errors would not be.

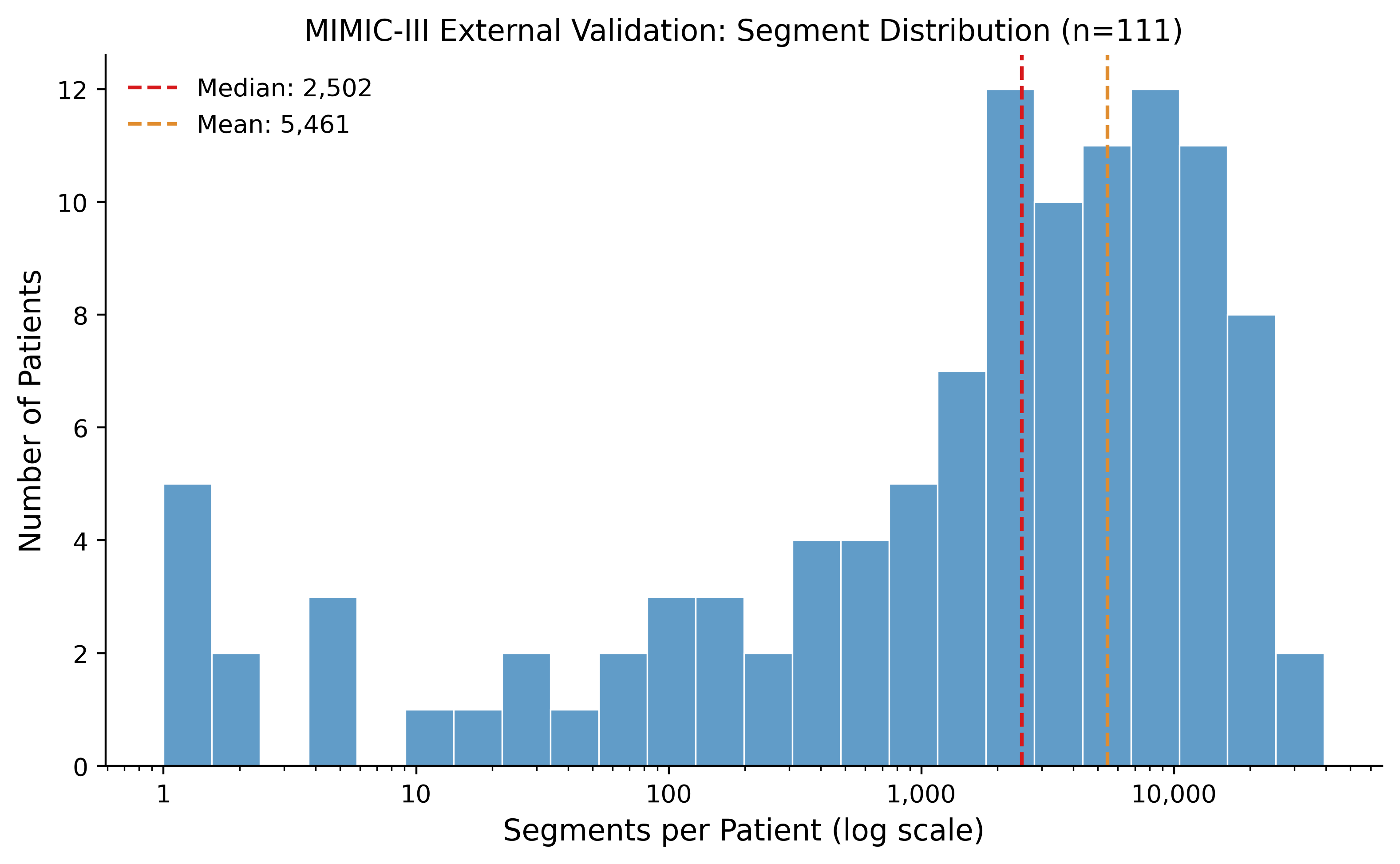

**Supplementary Fig. 1.** Segment count distribution per patient (MIMIC-III cohort). Log-scale histogram of segments per patient for MIMIC-III (external validation, n=111), with median and mean indicated. Log-scale was chosen to avoid imbalanced scaling caused by a few outlier patients.

**Analysis:** The distribution is right-skewed, with most patients contributing a moderate-to-large number of segments and a smaller number contributing very few. Eighteen patients (16.2% of the cohort) contributed fewer than 100 segments, comprising less than 0.1% of total external validation segments; a sensitivity analysis excluding these patients showed negligible change in performance across all model configurations (max ΔMAE = 0.0009 mmHg, max Δr = 0.0002), supporting patient-level bootstrapping as an appropriate mitigation for this imbalance.

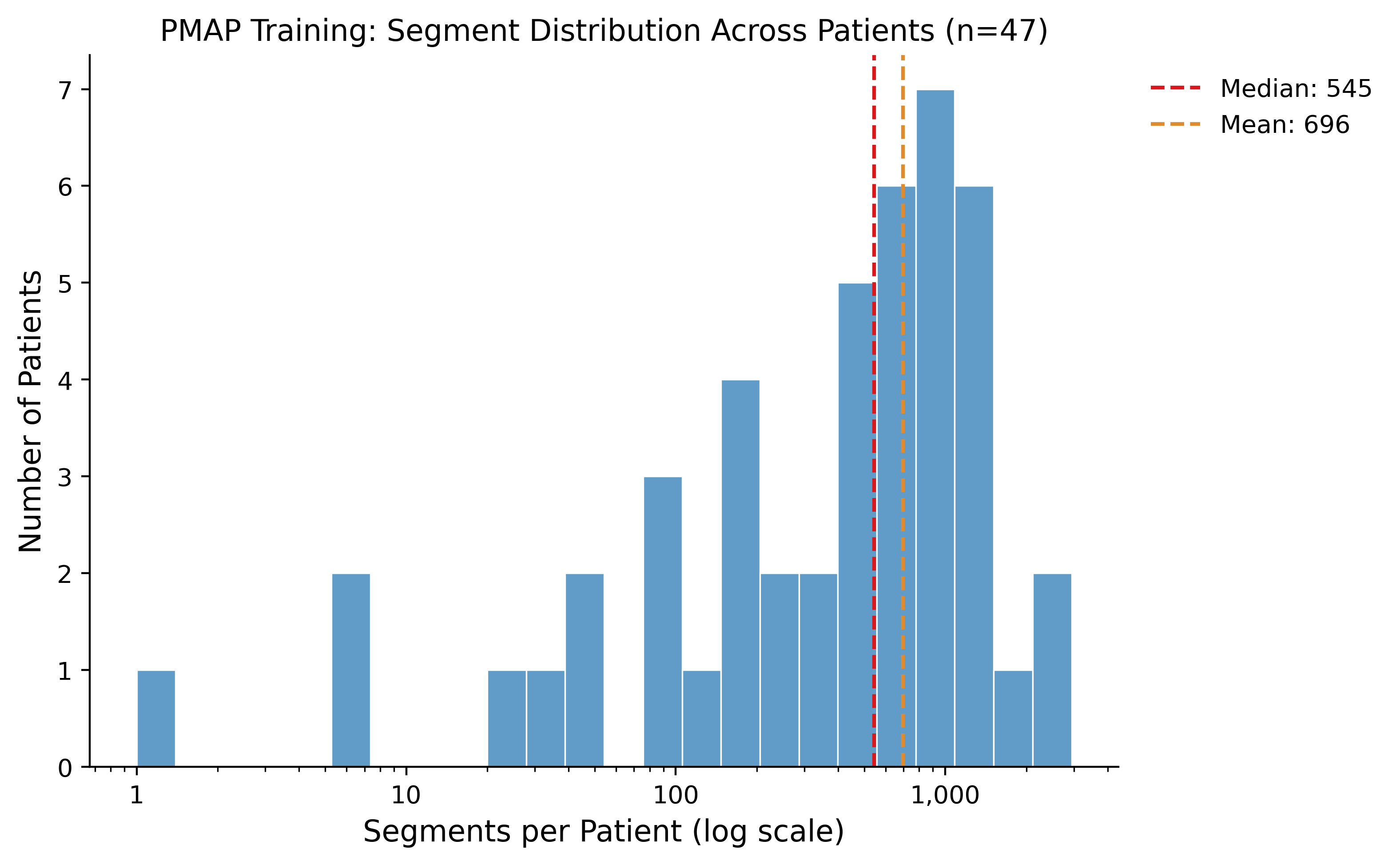

**Supplementary Fig. 2.** Segment count distribution per patient (PMAP cohort). Log-scale histogram of segments per patient for PMAP (training/internal validation, n=47), with median and mean indicated. Log-scale was chosen to avoid imbalanced scaling caused by a few outlier patients.

**Analysis:** PMAP showed a similarly right-skewed distribution to MIMIC-III (Supplementary Fig. 1). Given the smaller overall training set, patients contributing few segments were retained rather than excluded to maximize available training data.

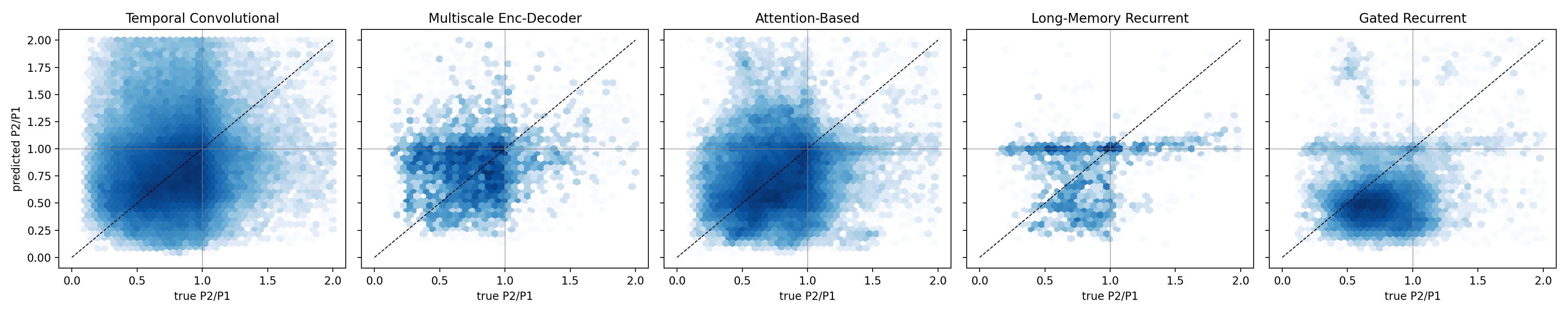

**Supplementary Fig. 3.** Predicted versus reference P2/P1 amplitude ratio, internally selected configurations (MIMIC-III external validation; 10,030 segments, 111 patients). Hexagonal density plots of beat-level P2/P1 ratios for beats in which P1 and P2 were detected in both the reference and predicted waveform; dashed line indicates identity. The multiscale encoder-decoder and long-memory recurrent models contributed few beats with a detected P1–P2 pair (see Supplementary Table 17), and predicted ratios cluster near 1 or fall in narrow bands irrespective of the reference value.

**Analysis:** Predicted P2/P1 ratios show little relationship to the reference ratio for any architecture (Supplementary Table 17: r = 0.14–0.40), and the models regress toward a typical ratio rather than tracking beat-to-beat variation in peak configuration. This is consistent with the representative waveforms in Figs. 3 and 4, in which several models produce similar smoothed pulse shapes across P1-dominant and P2-dominant beats.
